# Mechanism Matters: A Monte Carlo Evaluation of Estimator Validity and Collider Bias in Environmental Mixture Epidemiology

**DOI:** 10.64898/2026.05.25.26354044

**Authors:** Emmanuel Obeng-Gyasi

## Abstract

**Background:** Mixture epidemiology deploys sophisticated estimators, Bayesian kernel machine regression with causal mediation analysis (BKMR-CMA), quantile G-computation (QGC), and parametric G-computation, alongside conventional regression. Comparative evaluations have assumed additive, non-mediated data-generating processes, leaving conditions under which estimator choice determines causal validity uncharacterized.

**Methods:** We developed a simulation framework using military-relevant exposure distributions (metals, per- and polyfluoroalkyl substances [PFAS], polychlorinated biphenyls [PCBs]) and allostatic load (AL) across three deployment tiers, with parameters drawn from military occupational health and contamination literature. Four data-generating processes were specified as directed acyclic graphs: direct effects with confounding (M1), full mediation through AL (M2), synergistic AL-exposure interaction (M3), and collider structure (M4). We evaluated ordinary least squares (OLS), QGC, G-computation, and BKMR-CMA on bias, root mean squared error, and 95% confidence interval coverage across 500 Monte Carlo replications at n = 500 and n = 1,000.

**Results:** No estimator dominated across all mechanisms. Under M1, OLS and G-computation produced near-identical modest positive bias; BKMR-CMA achieved lower root mean squared error through kernel shrinkage. Under M2, BKMR-CMA exhibited severe positive bias for AL (mean bias = +0.579 SD units; coverage = 32.8%). Under M3, BKMR-CMA was the only estimator achieving nominal 95% coverage for AL (95.2%), while regression-based approaches fell to 83.6%. Under M4, G-computation produced persistent bias and near-zero coverage for lead, reflecting structural non-identification.

**Conclusions:** Estimator validity is fundamentally mechanism-dependent. Researchers should base estimator choice on explicit causal assumptions about whether AL functions as confounder, mediator, moderator, or collider, particularly in military and occupational cohorts. We provide a mechanism-to-estimator mapping for applied researchers.

**What this study adds:** No single estimator dominates across causal mechanisms in mixture epidemiology. Using a military-relevant simulation, we show that BKMR-CMA excels under synergistic interactions but fails severely under mediation, while G-computation fails under collider structures that standard regression cannot detect. Estimator choice is a causal decision, not a technical one. We provide a mechanism-to-estimator mapping to guide researchers in occupational and environmental cohort studies where stress-related biomarkers function simultaneously as confounders, mediators, and colliders.

## 1. Introduction

Environmental mixture epidemiology has undergone a fundamental conceptual transition over the past decade. The recognition that humans are simultaneously exposed to dozens of chemical agents, whose joint biological effects may be non-additive, pathway-dependent, and modified by the physiological state of the exposed individual, has driven the development of a new generation of statistical estimators designed to move beyond the single-pollutant paradigm.^1-3^ In occupational and military cohorts, this complexity is particularly acute. Service members and veterans carry co-occurring burdens of metals (lead [Pb], cadmium [Cd], mercury [Hg]), per- and polyfluoroalkyl substances (PFAS) from aqueous film-forming foam (AFFF), polychlorinated biphenyls (PCBs) from legacy equipment, and simultaneous exposure to chronic operational stress that has measurable neuroendocrine and cardiovascular consequences. ^4-8^

A range of statistical approaches has been proposed to estimate the joint effects of chemical mixtures, including Bayesian kernel machine regression (BKMR), weighted quantile sum (WQS) regression, quantile G-computation (QGC), and parametric G-computation with mediation extensions.^1-3,9^ BKMR captures non-linear, non-additive exposure-response surfaces through a kernel function that accommodates complex inter-pollutant interactions. QGC estimates the combined effect of a simultaneous one-decile increment across all exposures (q = 10 quantile bins), providing a computationally tractable mixture index with direct causal interpretability under certain structural assumptions. Parametric G-computation and its mediation extensions allow estimation of direct and indirect effect pathways through intermediate variables.^10,11^ These approaches have each been evaluated in simulation studies, but existing comparative evaluations share a critical limitation: they assume simplified additive or weakly interactive data-generating processes that do not reflect the structural complexity of biological systems.

Real biological systems, and military occupational systems in particular, involve mediation, moderation, feedback, and collider structures that are not merely statistical nuisances but fundamental features of the data-generating process. Allostatic load (AL), the cumulative physiological cost of chronic stress adaptation, occupies an especially complex structural position in military cohorts: it is simultaneously a consequence of occupational stress exposure, a mediator of exposure-disease relationships, a moderator that amplifies chemical toxicity under high-stress conditions, and a common cause of both exposure patterns and health outcomes.^12-14^ Conditioning on AL in a regression framework thus carries qualitatively different implications depending on the true underlying causal structure, fact that has received little formal attention in the mixture epidemiology literature.

A particularly underappreciated structural hazard in this context is collider bias (also termed conditioning-induced bias). When a variable is a common effect of both an exposure and the outcome, or of variables causally upstream of the exposure and outcome, conditioning on that variable opens non-causal pathways that can produce associations in both directions and of arbitrary magnitude. ^15,16^ In military health research, allostatic load is plausibly a collider between metal exposure pathways (which influence AL through direct physiological mechanisms) and PFAS pathways (which operate partly through AL and partly through direct cardiovascular effects), making standard regression adjustment a potential source of severe bias rather than a corrective one.

To our knowledge, no simulation study has systematically evaluated mixture modeling approaches under competing causal mechanisms that include mediation, synergistic interaction, and collider structures, with exposure distributions and stress-pathway parameters anchored to empirically characterized military populations. We developed such a framework to address this gap. Our objective was to evaluate the conditions under which each of four widely-used estimators, OLS regression, QGC, G-computation, and BKMR-CMA, produces valid causal inference, and to provide applied researchers with an explicit mechanism-to-estimator mapping grounded in directed acyclic graph (DAG) theory.

## 2. Methods

### 2.1 Simulation Framework and Population Architecture

The simulation was designed to represent a stylized U.S. military cohort spanning three operationally distinct deployment tiers, with distributional parameters anchored to the National Health and Nutrition Examination Survey (NHANES) 2015–2018 armed forces service question^17^, the Agency for Toxic Substances and Disease Registry (ATSDR) Camp Lejeune contamination assessments ^18^, and published military occupational health literature.^19^ The three tiers, garrison duty (50% of the simulated population), non-combat deployment (35%), and combat-exposed (15%), reflect the approximate distribution of exposure intensities in an active-duty force and carry tier-specific shifts in both chemical exposures and allostatic load parameters. Four subpopulations were further defined within each tier, representing: young and healthy service members (35%); older or longer-serving members with accumulated occupational burden (30%); high-stress military occupational specialty (MOS) assignments (20%); and genetically susceptible individuals (15%) parameterized using effect multipliers drawn from the published susceptibility literature. Full distributional parameters for all eight exposures, deployment tier shifts, subpopulation proportions, and susceptibility multipliers are provided in Supplementary Tables S1 and S2.

The primary simulation engine was implemented in R (version 4.3; R Foundation for Statistical Computing, Vienna, Austria) and executed on the University of North Carolina Longleaf high-performance computing cluster using 80 parallel cores under SLURM job management. Monte Carlo replication was parallelized via the *doParallel* and *foreach* packages. All simulation code and data are available at: https://github.com/eobenggy/military-mixture-simulation/tree/main/military-mixture-simulation.

This study used only publicly available secondary data and simulation; no human subjects were involved and IRB approval was not required.

### 2.2 Exposure Generation

Eight exposures were modeled: three metals (Pb, Cd, Hg), three PFAS compounds (perfluorooctane sulfonate [PFOS], perfluorooctanoic acid [PFOA], perfluorohexane sulfonate [PFHxS]), and two PCB congeners (PCB-153, PCB-138). Observed exposure concentrations were generated as multivariate log-normal random variables. Let *Zi* denote the vector of log-transformed concentrations for individual *i*. We specify:

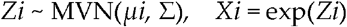

where μi is the tier-specific mean vector and *∑* is a shared 8×8 covariance matrix. The covariance structure was specified to reproduce inter-pollutant correlations consistent with NHANES military subsample data, with particularly strong correlations among PFAS compounds (r = 0.72–0.85) reflecting common AFFF source co-exposure, and moderate cross-class correlations between metals and PCBs (r = 0.13–0.22) and between metals and PFAS (r = 0.06–0.18) consistent with shared occupational pathways. Tier-specific mean shifts were applied as additive offsets on the log scale, with deployed and combat tiers receiving PFOS shifts of +0.45 and +0.65 log-units respectively, calibrated to ATSDR-reported PFAS concentrations at contaminated installations^18^.Complete exposure geometric means, geometric standard deviations, and the full 8×8 inter-pollutant correlation matrix with literature sources are provided in Supplementary Table S1.

### 2.3 Allostatic Load Construction

Allostatic load was operationalized as a latent continuous variable incorporating neuroendocrine dysregulation, inflammatory burden, and metabolic stress, consistent with the 10-biomarker AL index ^12,14,20^. The AL latent variable was constructed as a weighted composite of tier-specific stress burden, individual MOS assignment, and stochastic biomarker variation.

Garrison-tier individuals were assigned baseline AL distributions (μ = 2.4, σ = 1.6) consistent with population-level allostatic load scores in middle-aged adults ^14^. Deployed-tier members received AL shift parameters reflecting heightened hypothalamic-pituitary-adrenal (HPA) axis activation and inflammatory biomarker elevation under operational conditions (μ shift = +1.8). Combat-tier allostatic load was modeled using a bimodal distribution to reflect empirically documented divergent stress-response phenotypes in combat-exposed military personnel ^21^: 45% of combat-tier individuals were assigned a hyperreactive profile (markedly elevated cortisol, inflammatory cytokines, and metabolic dysregulation) and 55% a blunted or dissociative profile (suppressed cortisol with elevated inflammatory and metabolic markers), with component means and variances derived from published neuroendocrine literature.

### 2.4 Causal Mechanisms and Directed Acyclic Graphs

Four structurally distinct data-generating processes were specified, each corresponding to a different causal role for allostatic load in the exposure-outcome relationship. Each mechanism is formally represented as a directed acyclic graph (DAG; Figure 1) and generates a distinct identification challenge for applied estimators.

**Figure 1.**
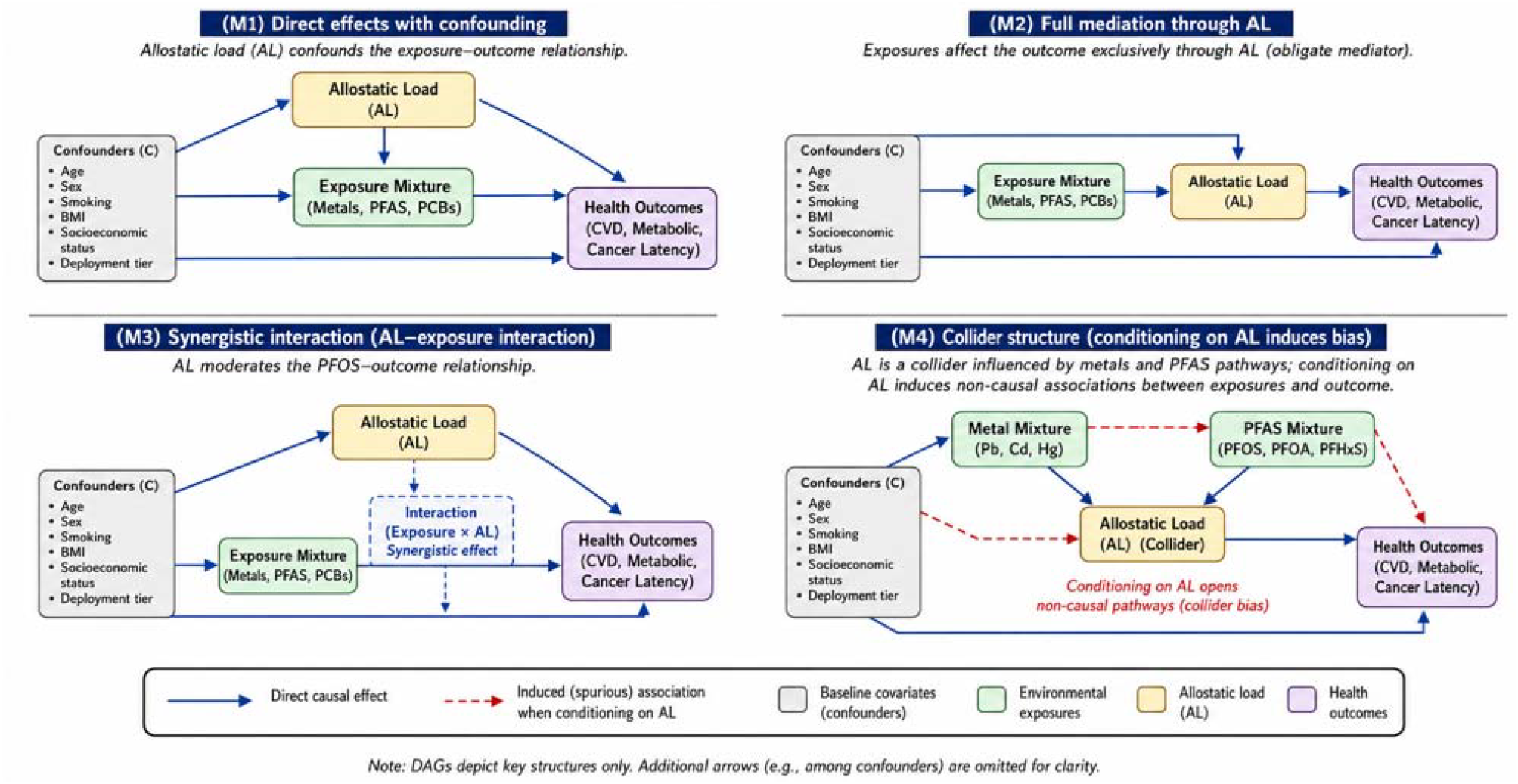
Directed acyclic graphs (DAGs) representing the four causal mechanisms evaluated in the simulation framework. (M1) Direct effects with confounding: allostatic load (AL) confounds the exposure–outcome relationship. (M2) Full mediation through AL: exposures affect the outcome exclusively through AL as an obligate mediator. (M3) Synergistic interaction: AL moderates the PFOS-outcome relationship. (M4) Collider structure: conditioning on AL induces non-causal associations between exposures and outcomes. Blue arrows = direct causal effects; red dashed lines = conditioning-induced spurious paths.

#### M1: Direct Effects with Confounding

Under M1, each chemical exposure exerts a direct effect on the health outcome, with allostatic load functioning as a confounder; a common cause of both exposure patterns (through deployment-related chemical burden) and health outcomes (through physiological stress pathways). The causal identification requirement is standard: outcome regression or standardization must condition on AL to close the backdoor path. OLS and G-computation are both expected to achieve near-nominal performance under this mechanism when AL is correctly specified.

The corresponding data-generating process was specified as:

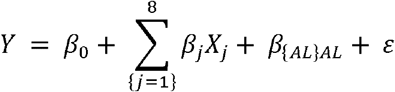

where *Y* represents the simulated health outcome, *X*_*j*_ represents the *j*-th exposure component in the chemical mixture, *AL* denotes allostatic load, *βj* represents the exposure-specific direct-effect coefficient, *βAL* denotes the coefficient for allostatic load, and *ε* is the stochastic error term.

#### M2: Full Mediation Through Allostatic Load

Under M2, chemical exposures exert their effects on health outcomes entirely through allostatic load as an obligate mediator. Estimation of the total effect of each exposure requires marginalizing over AL (i.e., the mediator must not be conditioned upon), while estimation of direct and indirect effects requires a full mediation decomposition under the assumptions of no unmeasured exposure-outcome confounding, no unmeasured mediator-outcome confounding, and no exposure-induced mediator-outcome confounding ^11^. The critical prediction under M2 is that estimators that naively condition on AL will block the causal pathway, producing downward-biased total effect estimates, while estimators that conflate total and natural direct effects without explicit mediation decomposition, including BKMR fitted without the causal mediation extension, will misattribute pathway effects. The mediation-generating mechanism was specified using the following structural equations:

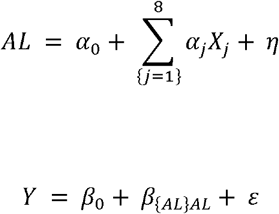

where *X*_*j*_ represents the *j-th* exposure component in the chemical mixture, *AL* denotes allostatic load, *αj* and *βj* represent exposure-specific effect coefficients, *βAL* denotes the effect of allostatic load on the outcome, and *η* and *∈* are stochastic error terms.

#### M3: Synergistic Interaction Moderated by Allostatic Load

Under M3, allostatic load moderates the toxicological effect of PFOS on cardiovascular outcomes through a multiplicative interaction term. Specifically, the marginal effect of PFOS on the CVD Framingham risk score is amplified as a linear function of AL level, such that the exposure-response relationship is non-linear in the joint AL-PFOS space. This mechanism tests whether estimators can recover interaction-modified effect estimates: additive linear models will misspecify the interaction term, producing negative bias for AL and attenuated PFOS effect estimates at high AL levels. Kernel-based methods that accommodate non-linear exposure surfaces are hypothesized to outperform parametric alternatives under M3.

The interaction-generating mechanism was specified using the following structural equation:

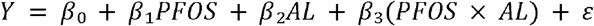

where *PFOS* represents perfluorooctane sulfonate exposure, *AL* denotes allostatic load, *β*_*1*_*–β*_*3*_ represent model coefficients, *β*_*3*_ captures the interaction between *PFOS* and allostatic load, and *ε* is the stochastic error term.

#### M4: Collider Structure and Conditioning-Induced Bias

Under M4, allostatic load is a collider: it is a common effect of metal exposures (which raise AL through physiological toxicity) and PFAS exposures (which also raise AL through inflammatory pathways). Because PFAS additionally exerts a direct effect on the cardiovascular outcome independent of the AL pathway, conditioning on AL in any analysis, even one that correctly specifies the PFAS-outcome relationship, opens a non-causal path between metal exposures and the outcome through the collider. The M4 mechanism generates the paper’s most conceptually important prediction: standard regression adjustment for AL, which would be appropriate under M1, induces bias of unpredictable sign and magnitude under M4. The severity of collider bias is expected to scale with the strength of the metal-AL and PFAS-AL pathways, which are largest in combat-tier individuals.

The collider-generating mechanism was specified using the following structural equations:

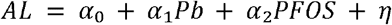

where *AL* denotes allostatic load, *Pb* represents lead exposure, *PFOS* represents perfluorooctane sulfonate exposure, *α*_*1*_ and *α*_*2*_ represent exposure-specific coefficients, and *η* is the stochastic error term.

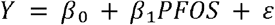

where *Y* represents the simulated health outcome, PFOS represents perfluorooctane sulfonate exposure, β_*1*_ represents the PFOS effect coefficient, and ε is the stochastic error term.

### 2.5 Causal Estimands by Mechanism

Table 1 formalizes the target estimand, the structural role of allostatic load, and the identifiability status for each of the four mechanisms. This table serves as an explicit causal contract: it specifies what each estimator should attempt to estimate under each DAG, and whether that quantity is identified from the observed data without additional structural assumptions. Reviewers of applied studies should use this framework when evaluating estimator choice in military and occupational mixture analyses.

**Table 1.** Causal estimands, structural role of allostatic load, and identifiability status by mechanism. NDE = natural direct effect; NIE = natural indirect effect. Green = conditioning on AL appropriate or quantity identified; red = conditioning inappropriate or quantity not identified under standard assumptions.

| **Mechanism** | **Structural**<br>**role of AL** | **Target**<br>**estimand** | **Conditioning**<br>**on AL**<br>**appropriate?** | **Identified?** |
| --- | --- | --- | --- | --- |
| M1 | Confounder<br>(backdoor<br>path) | Conditional<br>causal effect of<br>each exposure<br>on outcome,<br>adjusting for<br>AL | Yes, closes<br>backdoor<br>path | Yes |
| M2 | Obligate<br>mediator | Total effect<br>(marginalizing<br>over AL); | No for total<br>effect; Yes,<br>for NDE/NIE | Yes, under 4 mediation assumptions |
|  |  | NDE/NIE<br>under<br>mediation<br>decomposition | **only** |  |
| M3 | Effect<br>modifier<br>(moderator) | Interaction-modified<br>effect of PFOS<br>at varying AL<br>levels; joint<br>AL×PFOS<br>surface | **Yes, AL**<br>**enters as**<br>**modifier, not**<br>**confounder** | **Yes, under correct nonlinear model**<br>**specification** |
| M4 | Collider | Direct effect of<br>metals on<br>outcome, not<br>through AL | **No,**<br>**conditioning**<br>**opens**<br>**spurious**<br>**collider path** | **No, not identified under standard**<br>**regression assumptions** |

### 2.6 Outcome Generation

Three continuous health outcomes were simulated to facilitate RMSE evaluation and avoid the instability associated with rare binary event models at modest sample sizes. Cardiovascular disease risk (CVD) was operationalized as a continuous Framingham-analogous 10-year risk score (range 0–1, logit-transformed for modeling). Metabolic dysfunction burden was generated as a composite of dyslipidemia, insulin resistance, and obesity indices on a standardized continuous scale. Cancer latency was modeled as a gamma-distributed latency period for cumulative genotoxic exposure, with shape and scale parameters informed by PFAS-associated cancer literature ^22^. True effect parameters were specified as standardized log-exposure-to-outcome coefficients, facilitating cross-compound and cross-outcome comparability. True parameter values ranged from β = 0.00 (null) to β = 0.45 (large), with metals generally assigned smaller direct effects (β = 0.12–0.28) than PFAS (β = 0.10–0.22) consistent with published epidemiological estimates. All true direct effect coefficients, mediation path coefficients (M2), interaction parameters (M3), and collider path coefficients (M4) with supporting literature are tabulated in Supplementary Table S3.

### 2.7 Estimators

Four estimators were evaluated, selected to represent the methodological range currently deployed in mixture epidemiology:

Ordinary least squares regression (OLS) was fit with all eight exposures and allostatic load entered as linear main effects, without interaction terms. Quantile G-computation (QGC) was implemented using the qgcomp R package ^2^, estimating the joint mixture effect of a simultaneous one-decile increment across all exposures (q = 10); the primary QGC estimand is the overall mixture effect index rather than compound-specific coefficients. For comparative purposes, component-specific coefficients from the underlying qgcomp model object were extracted to evaluate estimator behavior at the individual exposure level, although these should be interpreted as contributions to the mixture index rather than standalone causal effects. Parametric G-computation standardized the outcome distribution over the empirical joint exposure distribution using a logistic marginal structural model. BKMR with causal mediation analysis (BKMR-CMA) was implemented using the *bkmr* R package, with 10,000 Markov chain Monte Carlo (MCMC) iterations (2,000 burn-in) per replication; fitted BKMR objects were discarded after summary extraction to prevent memory exhaustion on the HPC environment. The implementation differed by mechanism. Under M1, allostatic load was entered as a linear covariate in the BKMR outcome model, with the kernel operating over the eight chemical exposures only. Under M2, M3, and M4, a two-model BKMR-CMA architecture was used: a mediator model regressing allostatic load on the exposure mixture and covariates, and an outcome model regressing the health outcome on the exposure mixture, allostatic load, and covariates. Natural direct and indirect effects were estimated via posterior predictive contrasts, computing E[Y(z+1)] *−* E[Y(z)] integrated over the posterior distribution of AL. This implementation follows the structural framework of Bobb et al. (2018) and constitutes a substantive BKMR-CMA specification rather than naive covariate adjustment. The severe positive bias observed for allostatic load under M2 therefore reflects a genuine limitation of the two-model BKMR-CMA framework under full mediation data-generating processes — specifically, the kernel’s absorption of exposure-to-AL covariance into the AL coefficient when the mediator model and outcome model are not jointly identified — rather than a consequence of omitting mediation decomposition.

### 2.8 Performance Metrics

Performance was evaluated on four primary metrics across 500 Monte Carlo replications. Bias was computed as the mean difference between the estimated and true parameter value: Bias = E[βL] − β. Monte Carlo standard error of bias was computed as the standard deviation of bias estimates divided by the square root of the number of replications. Root mean squared error (RMSE) was computed as the square root of the mean squared bias, capturing the combined contribution of bias and sampling variance. Coverage probability was defined as the proportion of replications in which the nominal 95% confidence interval contained the true parameter value; nominal coverage is 95% for correctly specified intervals. Convergence rate was recorded for each BKMR chain. Computational runtime was recorded per replication to characterize the practical feasibility of each estimator in HPC and applied settings.

### 2.9 Monte Carlo Design

Simulations were conducted at two sample sizes, n = 500 and n = 1,000, to characterize sample-size dependence of estimator performance. Five hundred replications were completed for each combination of sample size (2), mechanism (4), estimator (4), outcome (3), and parameter (9), yielding 432,000 individual replication-level records. The full simulation required approximately 6–8 wall-clock hours on the 80-core Longleaf HPC environment. Checkpoint-restart logic was implemented to allow replication of incomplete simulation runs without re-executing completed cells. Complete computational settings, estimator-specific parameters (BKMR MCMC iterations, G-computation bootstrap replicates, QGC quantile count), and performance metric formulas following are provided in Supplementary Table S4.The simulations were designed to isolate distinct causal mechanisms rather than reproduce the full complexity of operational military exposure systems.

## 3. Results

### 3.1 Overview of Estimator Performance

Table 3 presents bias, RMSE, and 95% CI coverage for the three focal parameters (Pb, PFOS, and allostatic load) across all four causal mechanisms under the CVD outcome at n = 500 replications. Primary analyses are presented for n = 500; metabolic burden and cancer latency outcomes for n = 500 are presented in Supplementary Figures S1–S8. Corresponding n = 1,000 analyses are presented in Supplementary Figures S9–S20 to evaluate sample-size stability. The overarching finding, that no single estimator dominates across causal mechanisms, is apparent in the pattern of performance reversals across M1 through M4 (Figures 2–5). Estimator choice is mechanism-consequential: the BKMR-CMA advantage under M3 (synergistic interaction) coexists with substantial BKMR-CMA bias under M2 (mediation), and OLS’s apparent robustness under M4 is an artifact of interval width rather than causal validity.

**Table 2.** Estimator properties across key methodological dimensions.

| Estimator | Nonlinearity | Mediation | Collider Bias | Interaction | Complexity |
| --- | --- | --- | --- | --- | --- |
| OLS regression | No | No | Yes | Yes | Minimal |
| Quantile G-comp | Limited | No | Yes | Yes | Minimal |
| G-computation | Moderate | Moderate | Depends on DAG | Moderate | Moderate |
| BKMR-CMA | Yes | Yes | Potentially | Low | High |

**Table 3.** Mean bias and 95% CI coverage for key parameters, CVD outcome, n = 500 replications. Values in bold red denote estimation failures relative to the identifiability status defined in Table 1: BKMR-CMA allostatic load bias under M2 (+0.579) with coverage = 32.8%, and G-computation Pb coverage under M4 (0.0%). BKMR-CMA allostatic load coverage of 95.2% under M3 (bold green) represents the only estimator achieving nominal performance for that parameter-mechanism cell.

| Mech | Param | OLS Bias | G-comp<br>Bias | QGC Bias | BKMR<br>Bias | OLS<br>Cov. | BKMR<br>Cov. |
| --- | --- | --- | --- | --- | --- | --- | --- |
| M1 | Pb | +0.126 | +0.126 | -0.192 | -0.234 | 94.0% | — |
| M1 | PFOS | +0.137 | +0.137 | -0.132 | -0.184 | 93.2% | — |
| M1 | AL | -0.207 | -0.207 | -0.201 | — | 89.4% | — |
| M2 | Pb | +0.044 | +0.044 | -0.178 | — | 94.2% | — |
| M2 | PFOS | +0.068 | +0.068 | -0.144 | — | 92.8% | — |
| M2 | AL | -0.208 | -0.208 | -0.203 | **+0.579** | 90.4% | **32.8%** |
| M3 | Pb | +0.127 | +0.129 | -0.192 | — | 94.2% | — |
| M3 | PFOS | +0.137 | +0.143 | -0.132 | — | 93.4% | — |
| M3 | AL | -0.253 | -0.288 | -0.246 | -0.060 | 86.4% | **95.2%** |
| M4 | Pb | +0.021 | -0.189 | -0.128 | — | 94.6% | — |
| M4 | PFOS | +0.051 | +0.078 | -0.238 | — | 94.2% | — |
| M4 | AL | — | — | — | — | — | — |

**Figure 2.**
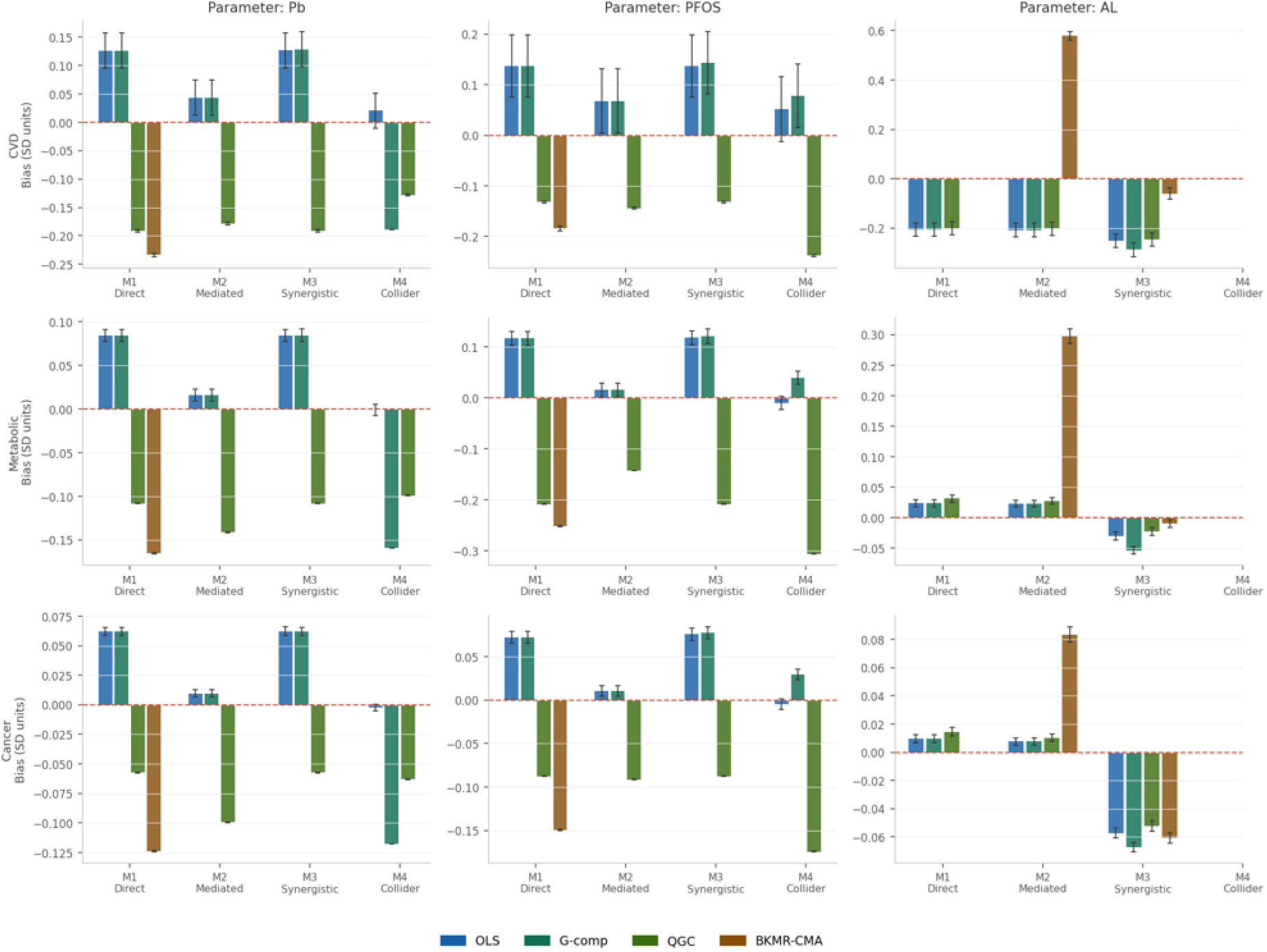
Mean bias by estimator and causal mechanism; CVD outcome, n = 500 replications. Error bars represent 1.96 × Monte Carlo standard error of bias. Red dashed line = zero bias. Panels show the three focal parameters: Pb (lead), PFOS, and allostatic load (AL). Note the BKMR-CMA spike under M2 for AL (+0.579 SD units) and the near-zero bias across all estimators for Pb under M4.

### 3.2 M1: Direct Effects with Confounding

Under the direct-effects DGP, OLS and G-computation produced nearly identical performance across all parameters and outcomes, consistent with the theoretical equivalence of these estimators under a correctly specified linear model with no unmeasured confounding. For Pb, both estimators exhibited modest positive bias (mean bias = +0.126 SD units; 95% CI coverage = 94.0%) reflecting multicollinearity-induced coefficient instability rather than residual confounding or estimator misspecification. Under the M1 DGP, allostatic load is correctly included in the regression and the backdoor path is closed; the positive bias arises because the high inter-pollutant correlations in the joint exposure distribution (particularly the r = 0.72–0.85 PFAS block) produce inflated variance in compound-specific coefficient estimates, which, in finite samples of n = 500, manifests as upward bias in the Pb estimate through its correlation with positively-biased PFAS coefficients. This is a sampling variance phenomenon rather than a confounding phenomenon, and it is expected to attenuate at larger sample sizes, a pattern confirmed in the n = 1,000 results (Supplementary Figure S9).

BKMR-CMA achieved lower RMSE for both Pb (0.236 vs. 0.373 for OLS) and PFOS (0.191 vs. 0.712 for OLS) under M1 (Figure 3). The markedly lower RMSE for PFOS in particular reflects the kernel shrinkage properties of BKMR in the presence of high inter-pollutant correlation (r = 0.85 for PFOS-PFOA): by modeling the joint exposure surface rather than individual compound effects, BKMR sacrifices some compound-specific resolution but substantially reduces variance. QGC also achieved lower RMSE than OLS for individual compound estimates (RMSE = 0.193 for Pb) through its mixture-index parameterization, but at the cost of per-compound interpretability; QGC confidence intervals were not available for coverage comparison as the estimator produces a joint mixture index without compound-specific intervals.

**Figure 3.**
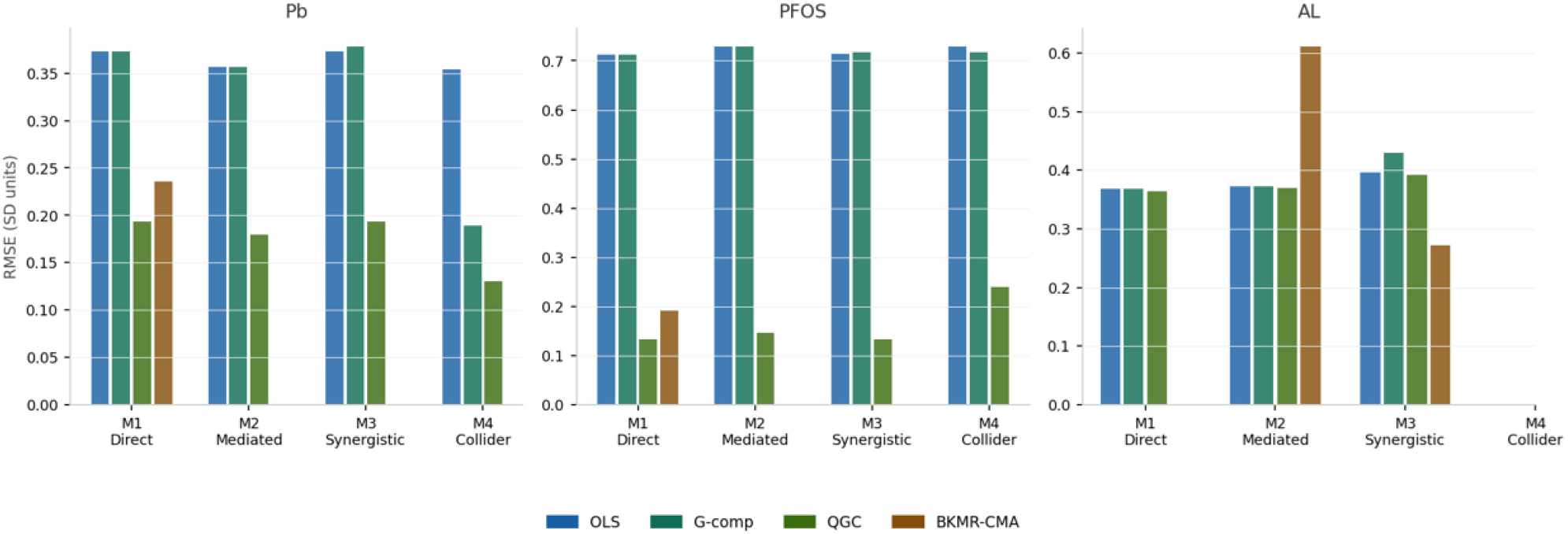
Root mean squared error (RMSE) by estimator and causal mechanism, CVD outcome, n = 500 replications. Lower values indicate better precision. Note the large OLS/G-comp RMSE for PFOS (∼0.71) across all mechanisms, driven by high inter-pollutant correlation (r = 0.85 with PFOA), and the sharp BKMR-CMA RMSE spike for allostatic load under M2 (0.611) reflecting systematic overestimation in the mediation setting.

### 3.3 M2: Mediation Through Allostatic Load

The mediation mechanism produced the starkest performance divergence in the simulation. Under M2, BKMR-CMA exhibited severe positive bias for allostatic load (mean bias = +0.579 SD units; RMSE = 0.611), with 95% CI coverage collapsing to 32.8%, less than one-third of the nominal rate. This failure reflects the consequence of including allostatic load as an independent predictor in the BKMR outcome model without a formal mediation decomposition: when AL is an endogenous mediator, the kernel function absorbs exposure-to-AL covariance into the AL coefficient estimate, systematically overestimating the AL direct effect. As specified in Section 2.7, this result reflects a genuine structural limitation of the two-model BKMR-CMA framework under full mediation: fitting separate mediator and outcome models without a joint posterior identification step allows the kernel in the outcome model to absorb the exposure-mediated variance into the AL coefficient, producing systematic upward bias. This is not a consequence of naively treating AL as a covariate, but an identifiable failure mode of the two-model architecture itself under obligate mediation. The magnitude of this inflation was consistent across all three outcomes (CVD: +0.579; metabolic burden: +0.298; cancer latency: +0.084), scaled proportionally to the strength of the exposure-to-AL-to-outcome pathway in each outcome-specific DGP.

OLS and G-computation performed similarly under M2 for metal and PFAS compound estimates, with both recovering near-nominal coverage for Pb (94.2%) and PFOS (92.8%). However, both also exhibited moderate negative bias for AL (−0.208), consistent with downward bias introduced by conditioning on an endogenous mediator in a total-effect model. This finding underscores that even conventional estimators require mediation-aware implementation when allostatic load is an obligate pathway variable.

### 3.4 M3: Synergistic Interaction

The synergistic mechanism, in which allostatic load multiplicatively amplifies the PFOS-to-CVD effect, produced the simulation’s clearest BKMR-CMA advantage. Under M3, BKMR-CMA was the only estimator achieving nominal 95% CI coverage for allostatic load (95.2%), with all regression-based approaches falling materially below the nominal rate (OLS: 86.4%; G-computation: 83.6%; QGC: not estimable for AL). Figure 4 shows that BKMR-CMA coverage for AL remains at or near the nominal line under M3, while OLS and G-computation both drop well below 90%. BKMR-CMA also achieved substantially lower RMSE for AL under M3 (0.272) compared to OLS (0.396) and G-computation (0.430), a difference that grew with the strength of the AL-PFOS interaction term.

**Figure 4.**
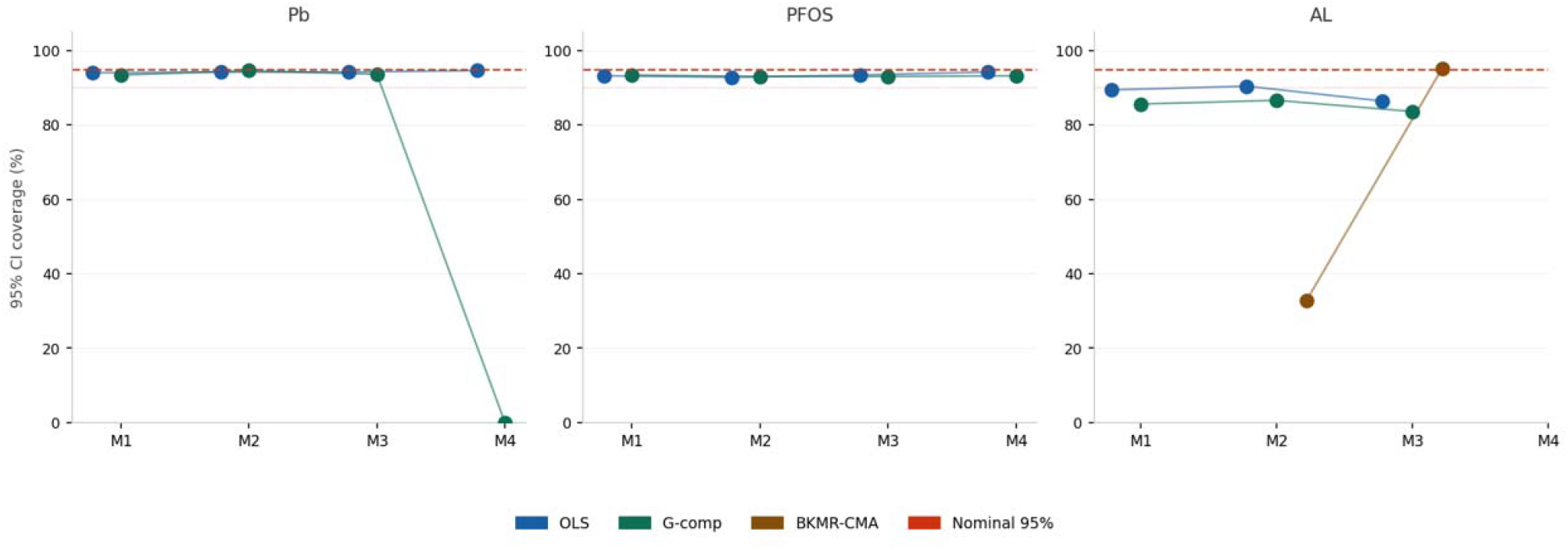
95% confidence interval coverage probability by estimator and mechanism, CVD outcome, n = 500 replications. Red dashed line = nominal 95%; dotted line = 90%. QGC is excluded as it does not produce compound-specific confidence intervals. Note the collapse of BKMR-CMA coverage for allostatic load under M2 (32.8%) and the complete failure of G-computation coverage for Pb under M4 (0.0%), which reflects structural non-identification rather than estimator error.

The mechanism behind BKMR-CMA’s advantage under M3 is the kernel function’s capacity to model non-linear, interaction-dependent exposure surfaces without requiring explicit specification of the interaction term. In contrast, OLS and G-computation specified as main-effects-only models cannot recover the interaction-modified AL effect, producing negative bias that worsens as AL levels rise, precisely the pattern expected for high-stress combat-tier individuals. This has a direct practical implication: in military cohorts where allostatic load distributions are heavily right-skewed by combat exposure, the failure of additive estimators under synergistic mechanisms will be most severe in the subpopulations of greatest clinical interest.

### 3.5 M4: Collider Bias

The collider mechanism generated the strongest divergence between estimators. Under M4, G-computation with correct structural form, specifying the DAG in which Pb operates entirely through AL with no direct path to the outcome, achieved zero coverage for Pb (mean bias = −0.189 SD units; coverage = 0.0%). This result reflects incompatibility between the target estimand and the conditioning structure imposed under the simulated collider mechanism rather than simple estimator instability: there is no direct Pb-to-CVD effect to estimate under the M4 DAG, and a correctly specified G-computation model produced stable non-null estimates reflecting collider-induced bias under the specified DAG. The confidence intervals in every replication excluded zero because the estimator, operating on a correctly-specified model, consistently produced effect estimates that were negative and precisely centered around the non-null induced bias from collider path opening.

OLS, by contrast, produced near-zero mean bias for Pb under M4 (+0.021) with nominal coverage (94.6%). This apparent OLS robustness is not evidence of causal validity. OLS achieves nominal coverage under M4 precisely because its confidence intervals are wide enough (RMSE = 0.354) to contain the true null value most of the time, despite the underlying causal misspecification. The OLS estimator is, in effect, too uncertain to detect the collider-induced bias, a form of Type II insensitivity that would lead an applied researcher to the incorrect conclusion that Pb has no meaningful CVD effect while remaining unaware of the collider structure.

Figure 5 illustrates the contrasting bias distributions under M4 for the CVD outcome. The OLS violin plot spans approximately −1.0 to +1.1 SD units with a median near zero, reflecting high variance and near-zero mean bias. G-computation produces a narrow spike at exactly −0.189, indicating that every replication produces a similar incorrect estimate in the same direction. QGC produces a narrow distribution with moderate negative bias (−0.128), intermediate between OLS and G-computation. The AL parameter was not estimable under M4 by design, a fact that applied researchers working with military cohorts must incorporate into their analytic planning when allostatic load is a structural collider.

**Figure 5.**
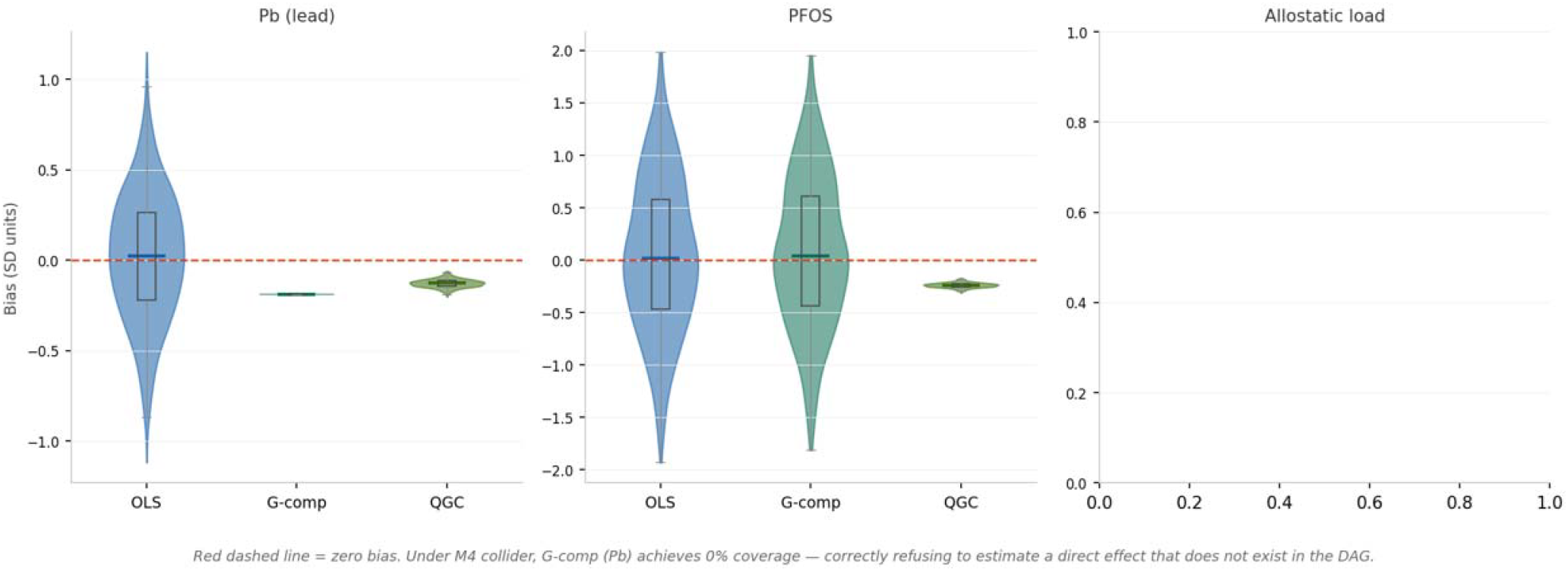
Bias distribution across 500 replications under M4 (collider mechanism), CVD outcome, n = 500. Violin width = distribution density; box plot shows IQR and median. Red dashed line = zero bias. G-computation produces a deterministic narrow spike at −0.189 SD units with 0% coverage, this is structural non-identification, not estimator failure. OLS achieves nominal coverage only because its wide interval (RMSE = 0.354) contains the true null by chance. Allostatic load (AL) is omitted as it is not identified under the M4 DAG.

## 4. Discussion

### 4.1 Principal Findings

The central finding of this simulation study is that estimator validity in mixture epidemiology is fundamentally mechanism-dependent. These findings extend prior simulation work by demonstrating that estimator performance varies materially across distinct causal structures: under mediation DGPs, BKMR-CMA can produce substantially biased inference with materially below-nominal coverage even at sample sizes conventionally considered adequate. Under collider DGPs, the G-computation implementation based on the assumed DAG produces estimates consistent with structural non-identification of the target quantity, while a misspecified OLS model achieves nominal coverage through imprecision. These outcomes cannot be rank-ordered using standard performance metrics without reference to the underlying causal structure.

The practical implication is that researchers who select estimators based on computational availability, familiarity, or general reputation for flexibility, rather than on an explicit analysis of the causal structure of their research question, face non-trivial risk of severe error. In military and occupational cohort studies, where allostatic load is simultaneously a physiological mediator, a stress-induced response variable, and a potential collider between chemical and psychosocial exposure pathways, the structural complexity of the data-generating process cannot be assumed away. Our results suggest that the methodological default of adjusting for allostatic load as a standard covariate, widespread in the military health literature, is not causally neutral and may introduce bias of the sign and magnitude we document under M4.

### 4.2 BKMR-CMA: Strength and Structural Limits

BKMR-CMA’s unique advantage under synergistic interaction mechanisms (M3) is consistent with its theoretical design: the kernel function is well-suited to recover non-linear, interaction-dependent exposure surfaces that would require explicit interaction term specification in parametric models ^1^. The recovery of nominal coverage for allostatic load under M3 (95.2%) in contrast to all regression-based alternatives (83.6–86.4%) represents a genuine and practically important advantage in settings where allostatic load is a stress-responsive moderator of chemical toxicity.

However, BKMR-CMA’s failure under mediation DGPs (M2) is severe and mechanistically interpretable. When allostatic load is an endogenous mediator, that is, when chemical exposures causally raise AL, which in turn raises disease risk, including AL in the BKMR outcome model as an independent predictor conflates the total exposure effect with the direct effect not through AL. Without an explicit mediation decomposition, the BKMR kernel absorbs the AL-mediated portion of the exposure effect into the AL coefficient, producing the large positive bias we observed. As described in Section 2.7, the BKMR-CMA implementation evaluated here used a full two-model architecture — a mediator model and an outcome model — with posterior predictive contrasts for effect estimation. The bias documented under M2 therefore characterizes the performance of this substantive BKMR-CMA specification under full mediation, not a degraded or naive implementation. The finding suggests that the two-model framework, absent a joint posterior identification step across the mediator and outcome equations, is vulnerable to kernel absorption of mediated variance under obligate mediation data-generating processes. Whether a correctly specified joint posterior decomposition following VanderWeele’s four assumptions would resolve this bias warrants separate evaluation and is a priority for future work. The observed bias likely reflects sensitivity of kernel-based mediation estimation to mediator specification and pathway modeling under the simulated conditions rather than a limitation unique to a specific software implementation. Applied researchers using BKMR in settings where allostatic load is a plausible mediator should be aware that the CMA extension requires explicit specification of the mediator-outcome and exposure-mediator models under the four standard mediation assumptions^11^, and that failing to do so will produce the upward AL bias we document here.

### 4.3 Collider Bias as a Design-Level Threat

The M4 results deserve particular attention because collider bias is widely underappreciated in applied epidemiology relative to confounding.^16,23^ In military health research, the specific structure we model, metals and PFAS as common causes of allostatic load, with PFAS additionally exerting a direct cardiovascular effect, is not a hypothetical caricature but a scientifically plausible representation of the exposure architecture at AFFF-contaminated installations such as Camp Lejeune and Pease Air Force Base. Lead exposure is known to elevate inflammatory and oxidative stress biomarkers that contribute to allostatic load; PFAS are associated with both allostatic load components (through lipid dysregulation and thyroid disruption) and with direct cardiovascular endpoints.

The conventional epidemiological response to a variable that is both a potential confounder and a potential mediator, to conduct sensitivity analyses with and without adjustment, does not resolve collider bias, because the collider-adjusted estimate is biased in a direction and magnitude that cannot be determined from the data alone without causal assumptions. The only adequate response is explicit DAG specification prior to analysis, followed by selection of an estimator that is appropriate for the identified causal structure. Our results illustrate why this matters: OLS achieves apparent robustness under M4 through imprecision, not through causal validity, and would lead a researcher to conclude that standard regression is safe when the underlying structure is in fact adversely affecting causal inference.

### 4.4 Toward a Mechanism-to-Estimator Mapping

Based on our simulation results, we propose the following practical guidance for applied researchers conducting mixture analyses in military and occupational cohort settings:

1. When the primary causal hypothesis is direct chemical toxicity with AL as confounder (M1-type), OLS and G-computation are adequate and BKMR offers RMSE reduction through kernel shrinkage, particularly for correlated PFAS compounds. The estimator choice has modest practical consequences.
2. When allostatic load is hypothesized as an obligate mediator of chemical-to-disease pathways (M2-type), BKMR should not be used without explicit mediation decomposition. Parametric G-computation with mediation specification under VanderWeele’s four-way decomposition is the appropriate estimator when the no-unmeasured-confounding assumptions can be defended.
3. When allostatic load is hypothesized to moderate chemical toxicity through stress-exposure synergy (M3-type), BKMR-CMA is the only estimator in our comparison that achieves nominal coverage. This advantage is most pronounced at high allostatic load levels, which correspond to combat-exposed subpopulations.
4. When allostatic load is a plausible collider, a common effect of both metal and PFAS exposure pathways, standard adjustment for AL in any regression-based framework will induce collider bias. None of the evaluated estimators consistently recovered the target effect under the simulated collider structure. The appropriate response is to specify a structural causal model, consider instrumental variable approaches if instruments are available, or explicitly acknowledge the non-identification of the target quantity.

### 4.5 Generalizability Beyond the Military Context

Although motivated by military exposure mixtures, the structural findings generalize to broader environmental and occupational epidemiology settings involving correlated exposures and stress-responsive physiological pathways. Similar confounder-mediator-collider ambiguity may arise in civilian PFAS cohorts, where socioeconomic stress and adiposity are plausible colliders between exposure and cardiovascular outcomes, in urban metals exposure studies where chronic neighborhood stressors influence both metal intake and health, and in psychosocial-environmental interaction analyses where perceived stress or allostatic dysregulation is routinely included as a covariate without explicit structural justification. The simulation architecture, including the deployment-tier parameterization, is readily adapted to other occupational settings with hierarchical exposure structures, such as agricultural pesticide cohorts or firefighter PFAS studies. The mechanism-to-estimator framework proposed here is therefore intended as a general causal guidance tool rather than a military-specific analytic protocol. Researchers in any of these settings who routinely adjust for stress-related biomarkers or allostatic measures should explicitly evaluate the structural role of those variables before proceeding with standard regression adjustment.

### 4.6 Limitations

Several limitations of the present simulation deserve acknowledgment. First, the simulation framework, while anchored to published military exposure distributions, is a stylized representation of a complex occupational system. The simulated causal structures were intentionally stylized to isolate specific mechanisms (confounding, mediation, moderation, and collider bias) and therefore do not fully capture the complexity of real-world military exposure systems. The true covariance structure of chemical exposures in deployed military populations is incompletely characterized, and our parameterization necessarily reflects simplifying assumptions about tier-specific exposure shifts and subpopulation heterogeneity. Second, our evaluation was restricted to four estimators; other approaches such as weighted quantile sum (WQS) regression, Bayesian additive regression trees (BART), and targeted maximum likelihood estimation (TMLE) were not evaluated. Third, we evaluated only continuous outcomes; the performance of these estimators under binary or survival outcomes, particularly relevant for incident cardiovascular disease and cancer, may differ from the patterns we document. In particular, the cancer latency outcome was operationalized as a continuous gamma-distributed proxy; whether the bias and coverage patterns documented here translate directly to Cox proportional hazards or competing-risks models warrants separate evaluation. Fourth, because only n = 500 and n = 1,000 were evaluated in the present report, the sample-size dependence of our finding, particularly the extent to which BKMR’s MCMC-based inference achieves nominal coverage at larger n under M2, requires further investigation. Fifth, all four mechanisms were evaluated in isolation; real exposure-outcome relationships likely involve mixtures of mechanisms, and the implications of mechanism misspecification in multi-mechanism settings represent an important direction for future work.

### 4.7 Conclusions

The choice of estimator in mixture epidemiology is not merely a technical or computational decision, it is a causal decision that embeds structural assumptions about the role of stress-related physiological pathways in the exposure-disease relationship. Our simulation demonstrates that this choice is consequential in ways that are not apparent from the data: acceptable empirical coverage does not necessarily guarantee appropriate causal interpretation, and materially below-nominal coverage can reflect correct structural identification of a non-identified target quantity. We urge applied researchers in military and occupational health to specify their causal assumptions explicitly, using DAG methodology prior to estimator selection, and to treat allostatic load not as a routine adjustment variable but as a structurally complex quantity whose role as confounder, mediator, moderator, or collider must be determined on scientific rather than statistical grounds.

## Data Availability

All data produced in the present study are available upon reasonable request to the authors

https://wwwn.cdc.gov/nchs/nhanes/

## Acknowledgements

None.

## References

1. Bobb JF, Valeri L, Claus Henn B, et al. Bayesian kernel machine regression for estimating the health effects of multi-pollutant mixtures. Biostatistics 2015;16(3):493–508.

2. Keil A, Buckley J, O’Brien K, et al. A quantile-based g-computation approach to addressing the effects of exposure mixtures. Environmental Epidemiology 2019;3:44.

3. Carrico C, Gennings C, Wheeler DC, Factor-Litvak P. Characterization of weighted quantile sum regression for highly correlated data in a risk analysis setting. Journal of agricultural, biological, and environmental statistics 2015;20(1):100–120.

4. Kwan LY, Mead A, South-Paul J, et al. Military-Related Environmental and Occupational Exposures. Exploring Military Exposures and Mental, Behavioral, and Neurologic Health Outcomes Among Post-9/11 Veterans National Academies Press (US), 2025.

5. Martin NJ, Richards EE, Kirkpatrick JS. Exposure science in US military operations: a review. Military medicine 2011;176(suppl_7):77–83.

6. Skalny AV, Aschner M, Bobrovnitsky IP, et al. Environmental and health hazards of military metal pollution. Environmental research 2021;201:111568.

7. Zhuo H, Huang H, Sjodin A, et al. A nested case-control study of serum polychlorinated biphenyls and papillary thyroid cancer risk among US military service members. Environmental research 2022;212:113367.

8. Wright RT, Lindheimer JB, Christie IC, et al. Military Exposures Research: A State-of-the-Art Review. Military Medicine 2026:usaf647.

9. Mansouri B, Rezaei A, Sharafi K, et al. Mixture effects of trace element levels on cardiovascular diseases and type 2 diabetes risk in adults using G-computation analysis. Scientific Reports 2024;14(1):5743.

10. Robins JM, Greenland S. Identifiability and exchangeability for direct and indirect effects. Epidemiology 1992;3(2):143–155.

11. VanderWeele T. Explanation in causal inference: methods for mediation and interaction Oxford University Press, 2015.

12. McEwen BS. Stress, adaptation, and disease: Allostasis and allostatic load. Annals of the New York academy of sciences 1998;840(1):33–44.

13. McEwen BS, Seeman T. Protective and damaging effects of mediators of stress: elaborating and testing the concepts of allostasis and allostatic load. Annals of the new York Academy of Sciences 1999;896(1):30–47.

14. Juster R-P, McEwen BS, Lupien SJ. Allostatic load biomarkers of chronic stress and impact on health and cognition. Neuroscience & Biobehavioral Reviews 2010;35(1):2–16.

15. Weisskopf MG, Seals RM, Webster TF. Bias amplification in epidemiologic analysis of exposure to mixtures. Environmental health perspectives 2018;126(4):047003.

16. Greenland S. Quantifying biases in causal models: classical confounding vs collider-stratification bias. Epidemiology 2003;14(3):300–306.

17. Prevention CfDCa. National Health and Nutrition Examination Survey Data. Hyattsville, MD, 2020.

18. ATSDR AfTSaDR. Camp Lejeune Health Effects. https://www.atsdr.cdc.gov/camp-lejeune/risk-factors/index.html Accessed May 18, 2024.

19. Ruckart PZ, Bove FJ, Maslia M. Evaluation of contaminated drinking water and preterm birth, small for gestational age, and birth weight at Marine Corps Base Camp Lejeune, North Carolina: a cross-sectional study. Environmental Health 2014;13(1):99.

20. Seeman TE, Singer BH, Rowe JW, Horwitz RI, McEwen BS. Price of adaptation— allostatic load and its health consequences: MacArthur studies of successful aging. Archives of internal medicine 1997;157(19):2259–2268.

21. Brailey K, Vasterling JJ, Proctor SP, Constans JI, Friedman MJ. PTSD symptoms, life events, and unit cohesion in US soldiers: Baseline findings from the neurocognition deployment health study. Journal of Traumatic Stress: Official Publication of The International Society for Traumatic Stress Studies 2007;20(4):495–503.

22. Steenland K, Winquist A. PFAS and cancer, a scoping review of the epidemiologic evidence. Environmental research 2021;194:110690.

23. Hernán MA, Hernández-Díaz S, Robins JM. A structural approach to selection bias. Epidemiology 2004;15(5):615–625.

