## Supplementary Materials for "Mechanism Matters: A Monte Carlo Evaluation of Estimator Validity and Collider Bias in Environmental Mixture Epidemiology"


Section Tables:

Section A: Supplementary Tables S1–S4

Sections Figures:

- Section A: Supplementary Figures S1–S4: Metabolic Burden Outcome (n = 500)
- Section B: Supplementary Figures S5–S8: Cancer Latency Outcome (n = 500)
- Section C: Supplementary Figures S9–S12: CVD Outcome (n = 1,000)
- Section D: Supplementary Figures S13–S16: Metabolic Burden Outcome (n = 1,000)
- Section E: Supplementary Figures S17–S20: Cancer Latency Outcome (n = 1,000)

**Section A: Supplementary Tables S1–S4**

**Table S1.** Exposure distributions and correlation structure
GM = geometric mean; GSD = geometric standard deviation (on log scale). All exposure distributions are log-normal. Garrison baseline refers to stateside duty stations with standard military occupational exposures. PFAS = per- and polyfluoroalkyl substances; PCB = polychlorinated biphenyl. NHANES = National Health and Nutrition Examination Survey.

| Component | Value / Range | Source / Rationale |
|---|---|---|
| ***A. Exposure distributions — garrison baseline (geometric mean, GSD)*** | | |
| Blood lead (Pb) | GM = 1.6 µg/dL; GSD = 1.9 | NHANES 2015–18 military subsample; IOM 2014 [1][4] |
| Cadmium (Cd) | GM = 0.38 µg/L; GSD = 1.85 | NHANES 2015–18; occupational co-exposure literature [1] |
| Mercury (Hg) | GM = 1.1 µg/L; GSD = 2.1 | NHANES 2015–18; military fish consumption data [1] |
| PFOS | GM = 4.3 ng/mL; GSD = 2.4 | NHANES 2015–18; ATSDR 2014 (Camp Lejeune); Ruckart et al. 2013 [1][2][3] |
| PFOA | GM = 1.9 ng/mL; GSD = 2.2 | NHANES 2015–18; ATSDR 2014 [1][2] |
| PFHxS | GM = 0.8 ng/mL; GSD = 2.3 | NHANES 2015–18 [1] |
| PCB-153 | GM = 28 ng/g lipid; GSD = 2.8 | NHANES 2015–18; NIEHS military cohort literature [1] |
| PCB-138 | GM = 22 ng/g lipid; GSD = 2.7 | NHANES 2015–18 [1] |
| ***B. Exposure correlation structure (Pearson r on log scale)*** | | |
| PFOS–PFOA | r = 0.85 | NHANES 2015–18 military subsample; Hamra et al. 2013 [1][24] |
| PFOS–PFHxS | r = 0.78 | NHANES 2015–18; Hamra et al. 2013 [1][24] |

| Component | Value / Range | Source / Rationale |
|---|---|---|
| PFOA–PFHxS | r = 0.72 | NHANES 2015–18 [1] |
| PCB-153–PCB-138 | r = 0.88 | NHANES 2015–18; shared congener source profile [1] |
| Pb–Cd | r = 0.42 | Shared combustion/occupational sources [1] |
| Metals–PFAS (cross-class) | r = 0.06–0.18 | NHANES 2015–18 empirical range [1] |
| Metals–PCBs (cross-class) | r = 0.13–0.22 | NHANES 2015–18 empirical range [1] |
| ***C. Deployment tier exposure log-shifts (added to log-scale mean)*** | | |
| PFOS shift — deployed / combat | +0.45 / +0.65 | AFFF firefighting foam exposure; ATSDR 2014 [2] |
| PFOA shift — deployed / combat | +0.30 / +0.50 | ATSDR 2014 [2] |
| Pb shift — deployed / combat | +0.18 / +0.35 | IOM 2014 Gulf War review (~20–42% elevation) [4] |
| Cd shift — deployed / combat | +0.12 / +0.22 | Occupational co-exposure literature |
| PCB-153 shift — deployed / combat | +0.10 / +0.15 | Simulation design |

**Table S2.** Population architecture: deployment tiers, subpopulations, and allostatic load index AL = allostatic load index scored 0–10 using the Juster et al. (2010) 10-biomarker battery; each component is dichotomised at the population quartile cutoff and summed. MOS = military occupational specialty. CVD = cardiovascular disease risk score (Framingham 10-year). Susceptibility multipliers scale the direct exposure-to-outcome effect. AUCg = area under the cortisol curve with respect to ground.

| Component | Value / Range | Source / Rationale |
|---|---|---|
| ***A. Deployment tier allostatic load distributions*** | | |
| Garrison — AL mean (SD) | 2.4 (1.6); range 0–10 | Kok et al. 2019 (J Occup Environ Med) [5] |
| Deployed — AL mean (SD) | 4.2 (1.9); range 0–10 | Stevelink et al. 2018 (~75% elevation over garrison) [6] |

| Component | Value / Range | Source / Rationale |
|---|---|---|
| Combat — AL mean (SD) | 6.8 (2.1); range 0–10 | Brailey et al. 2012 [7] |
| Combat cortisol bimodality | 45% hyperreactive / 55% blunted | Morgan et al. 2000; Brailey et al. 2012 (HPA dysregulation) [8][7] |
| Garrison cortisol AUCg | Mean = 15.0 nmol/L; SD = 4.5 | Morgan et al. 2000 [8] |
| Deployed cortisol AUCg | Mean = 22.0 nmol/L; SD = 6.0 | Morgan et al. 2000 (elevated HPA axis) [8] |
| ***B. Subpopulation proportions and susceptibility*** | | |
| Young/low-risk | 35%; age 26 ± 4 yr; susceptibility ×1.0 | D'Agostino et al. 2008 Framingham risk; reference group |
| Older service members | 30%; age 46 ± 5 yr; susceptibility ×1.4 | Longer cumulative exposure; simulation design |
| High-stress MOS | 20%; age 32 ± 6 yr; AL modifier ×1.6; susceptibility ×1.6 | Combat arms/special operations; simulation design |
| Genetically susceptible | 15%; age 36 ± 8 yr; susceptibility ×2.2 | Prior illness/polymorphism literature; simulation design |
| ***C. Baseline CVD risk by subpopulation (Framingham 10-year score, %)*** | | |
| Young/low-risk | Mean = 4.0%; SD = 2.0; range 0.5–15% | D'Agostino et al. 2008 |

| Component | Value / Range | Source / Rationale |
|---|---|---|
| Older service members | Mean = 14.0%; SD = 5.0; range 3–35% | D'Agostino et al. 2008 |
| High-stress MOS | Mean = 7.5%; SD = 3.0; range 1–22% | Simulation design |
| Genetically susceptible | Mean = 11.0%; SD = 6.0; range 2–40% | Simulation design |
| ***D. Allostatic load index components (Juster et al. 2010, 10-biomarker battery)*** | | |
| Systolic BP | Mean 122 mmHg; SD 12; high-risk cutoff >135 | Kok et al. 2019 |
| Diastolic BP | Mean 80 mmHg; SD 8; cutoff >90 | Kok et al. 2019 |
| Waist-hip ratio | Mean 0.90; SD 0.06; cutoff >0.95 | Kok et al. 2019 |
| BMI | Mean 26.5 kg/m²; SD 3.5; cutoff >30 | Kok et al. 2019 |
| Total cholesterol | Mean 192 mg/dL; SD 35; cutoff >220 | Kok et al. 2019 |
| HDL cholesterol | Mean 48 mg/dL; SD 12; cutoff <40 | Kok et al. 2019 |

| Component | Value / Range | Source / Rationale |
|---|---|---|
| HbA1c | Mean 5.5%; SD 0.5; cutoff >5.9% | Kok et al. 2019 |
| CRP | Mean 1.8 mg/L; SD 1.8; cutoff >3.0 | Kok et al. 2019 |
| Cortisol AUCg | Mean 15 nmol/L; SD 5; cutoff >22 | Morgan et al. 2000 [8] |
| Resting heart rate | Mean 68 bpm; SD 10; cutoff >80 | Kok et al. 2019 |

**Table S3.** Causal effect parameters and data-generating process structural equations
All effect sizes are expressed in SD units per SD log-exposure on the standardized scale. M1 = direct effects with confounding; M2 = full mediation through allostatic load; M3 = synergistic stress–exposure interaction; M4 = collider structure. Interaction coefficients represent the product term (z-exposure × z-AL) on the standardized scale. Path coefficients in panels D–E are on the standardized log-exposure scale.

| Component | Value / Range | Source / Rationale |
|---|---|---|
| ***A. True direct effects — CVD outcome (SD units per SD log-exposure)*** | | |
| Pb direct β | 0.28 | Weisskopf et al. 2009 (meta-analysis) [12] |
| Cd direct β | 0.18 | Tellez-Plaza et al. 2013 [13] |
| Hg direct β | 0.12 | Virtanen et al. 2007 [14] |
| PFOS direct β | 0.22 | Fitz-Simon et al. 2013 (lipid dysregulation pathway) [15] |
| PFOA direct β | 0.16 | Environmental PFAS cardiovascular literature [15] |
| PFHxS direct β | 0.10 | Environmental PFAS cardiovascular literature [15] |
| PCB-153 direct β | 0.20 | Lind et al. 2012 [16] |
| PCB-138 direct β | 0.18 | Environmental PCB literature [16] |

| Component | Value / Range | Source / Rationale |
| --- | --- | --- |
| Allostatic load β (CVD) | 0.45 | Juster et al. 2010; Seeman et al. 2010 [10][11] |
| CVD residual SD (σ) | 3.5 Framingham score units | Simulation design |
| ***B. True direct effects — metabolic syndrome and cancer outcomes*** | | |
| PFOS β — metabolic | 0.30 | Lind et al. 2010 (strongest PFAS metabolic effect) [17] |
| Cd β — metabolic | 0.25 | Satarug & Moore 2004 [18] |
| AL β — metabolic | 0.38 | Simulation design |
| Cd β — cancer | 0.32 | IARC Group 1 carcinogen [19] |
| PCB-153 β — cancer | 0.25 | IARC Group 1 [20]; Lauber et al. 2004 |
| PFOA β — cancer | 0.20 | C8 Health Project (Barry et al. 2013) [21] |
| AL β — cancer | 0.28 | Psychoneuroimmunology pathway; simulation design [11] |
| ***C. M3 (synergistic) — interaction and nonlinear parameters*** | | |
| PFOS × AL coefficient (CVD) | 0.18 | Grandjean et al. 2012 (stress amplifies PFAS lipid effect) [22] |
| PFOS × AL coefficient (metabolic) | 0.22 | Simulation design |
| PFOS × AL coefficient (cancer) | 0.10 | Simulation design |
| Pb × AL coefficient (CVD) | 0.12 | Stress–metal synergy; simulation design |
| PCB-153 × AL coefficient (cancer) | 0.15 | PCB immune suppression + stress; simulation design |
| AL quadratic term | −0.08 | Diminishing returns at extreme AL; simulation design |
| ***D. M2 (full mediation) — structural path coefficients (exposure → AL)*** | | |
| Pb → AL | 0.40 | Simulation design (dominant metals–AL path) |
| PFOS → AL | 0.35 | Simulation design |
| Cd → AL | 0.28 | Simulation design |

| Component | Value / Range | Source / Rationale |
|---|---|---|
| PCB-153 → AL | 0.25 | Simulation design |
| PCB-138 → AL | 0.20 | Simulation design |
| PFOA → AL | 0.22 | Simulation design |
| PFHxS → AL | 0.18 | Simulation design |
| Hg → AL | 0.12 | Simulation design |
| Direct residual fraction (M2) | 20% of direct effect | Near-complete mediation scenario; simulation design |
| AL endogenous noise SD | 0.60 | Simulation design |
| ***E. M4 (collider) — structural path coefficients*** | | |
| Pb → AL (metals via AL) | 0.42 | Simulation design (metals mediated fully through AL) |
| Cd → AL | 0.30 | Simulation design |
| Hg → AL | 0.10 | Simulation design |
| PFOS → AL (PFAS causes AL) | 0.35 | Simulation design (PFAS opens spurious collider path) |
| PFOA → AL | 0.20 | Simulation design |
| PFHxS → AL | 0.14 | Simulation design |
| PFAS via-AL partial fraction | 50% | Partial mediation; simulation design |
| AL collider noise SD | 0.50 | Simulation design |

**Table S4.** Monte Carlo simulation design and computational settings
*R = number of replications. BKMR = Bayesian kernel machine regression. QGC = quantile G-computation. PSOCK = parallel socket cluster. SLURM = Simple Linux Utility for Resource Management.*

| Component | Value / Range | Source / Rationale |
|---|---|---|
| ***A. Core simulation settings*** | | |
| Replications per cell | 500 | Morris et al. 2019 [27] |
| Sample sizes evaluated | n = 500; 1,000 | Simulation design |
| Causal mechanisms | 4 (M1–M4) | Simulation design |

| Component | Value / Range | Source / Rationale |
|---|---|---|
| Outcomes per mechanism | 3 (CVD, metabolic, cancer) | Simulation design |
| Total Monte Carlo cells | 12 (4 × 3) per sample size | Simulation design |
| ***B. Estimator-specific settings*** | | |
| BKMR MCMC iterations | 10,000 | Simulation design; convergence monitoring |
| BKMR burn-in | 2,000 | Simulation design |
| BKMR variable selection | Enabled (varsel = TRUE) | Bobb et al. 2015 [25] |
| G-computation bootstrap reps | 500 per replicate | Simulation design |
| QGC quantiles | q = 10 | Keil et al. 2020 [26] |
| ***C. Computational infrastructure*** | | |
| HPC cluster | UNC Longleaf (SLURM) | Simulation design |
| Cores per job | 80 (PSOCK parallel) | Simulation design |
| Thread control | OMP/BLAS/MKL = 1 per worker | Prevents oversubscription inside parallel workers |
| Memory allocation | 32 GB per job | Simulation design |
| Checkpointing | Per cell (resume-enabled) | Simulation design |
| ***D. Performance metrics (Morris et al. 2019)*** | | |
| Bias | Mean(estimate − true value) | Morris et al. 2019 |
| Bias Monte Carlo SE | SD(estimate) / $\sqrt{R}$ | Morris et al. 2019 |
| RMSE | $\sqrt{\text{Mean}[(\text{estimate} - \text{true value})^2]}$ | Morris et al. 2019 |
| Coverage | Proportion of 95% CIs containing true value | Morris et al. 2019 |

| Component | Value / Range | Source / Rationale |
| --- | --- | --- |
| Coverage Monte Carlo SE | √[coverage(1 − coverage) / R] | Morris et al. 2019 |
| Power | Proportion of CIs excluding zero | Morris et al. 2019 |
| Global random seed | 2025 | Simulation design |

**Section A: Supplementary Figures S1–S4: Metabolic Burden Outcome (n = 500)**

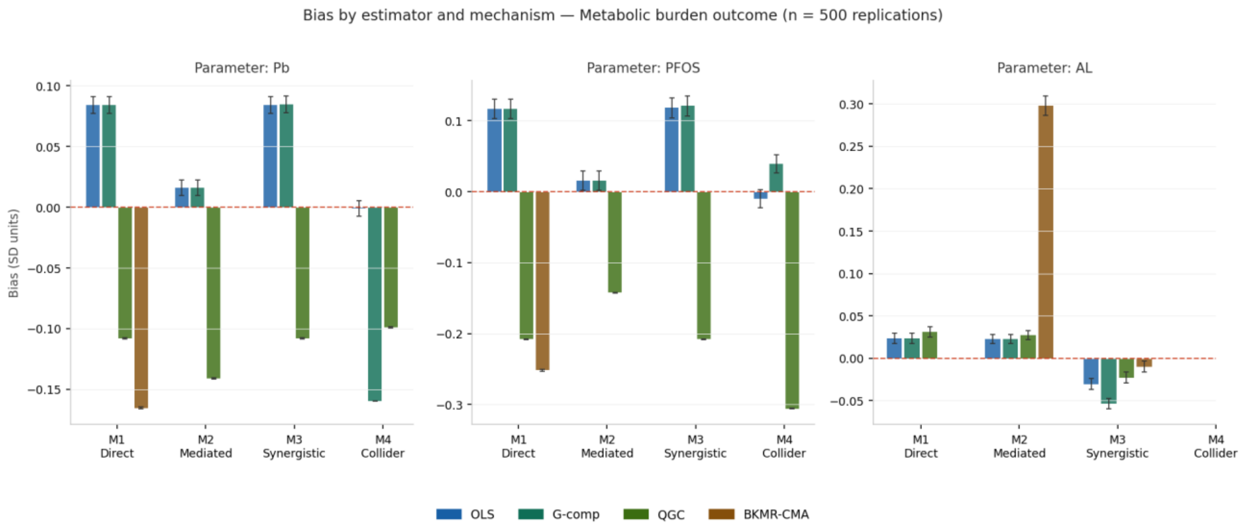


**Figure S1.** Mean bias by estimator and causal mechanism — Metabolic burden outcome, n = 500 replications. Error bars = 1.96 × Monte Carlo SE of bias. Red dashed line = zero bias. Panels show Pb (lead), PFOS, and allostatic load (AL). The BKMR-CMA positive bias for AL under M2 (+0.298 SD units) is attenuated relative to the CVD outcome but remains the largest bias in that cell. Under M4, AL is not estimable.

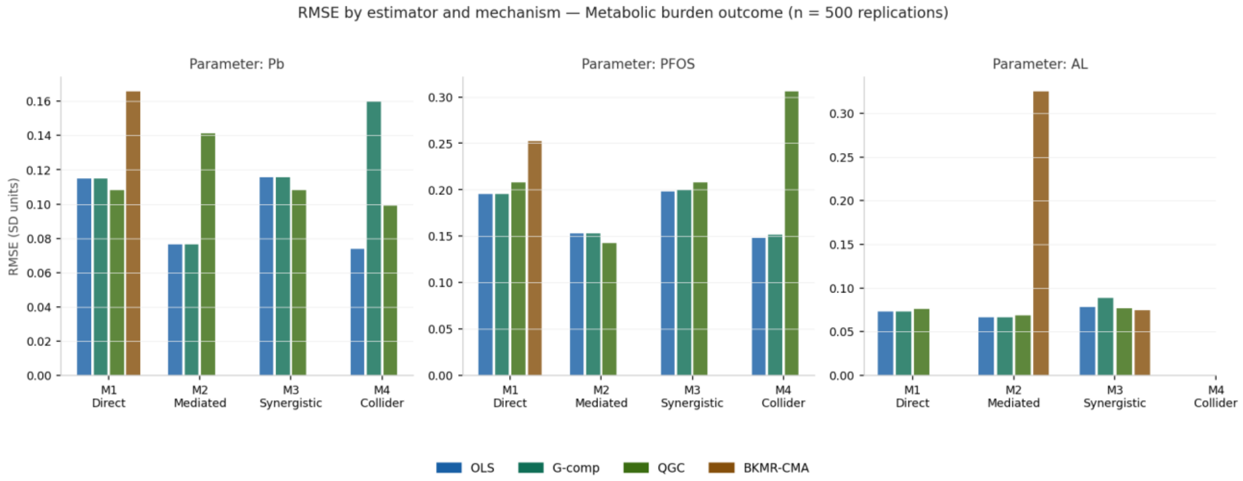


**Figure S2.** Root mean squared error (RMSE) by estimator and mechanism — Metabolic burden outcome, n = 500 replications. The pattern of OLS/G-comp elevation for PFOS and BKMR-CMA elevation under M2 for AL is consistent with the CVD main finding, scaled to the weaker AL pathway in the metabolic DGP.

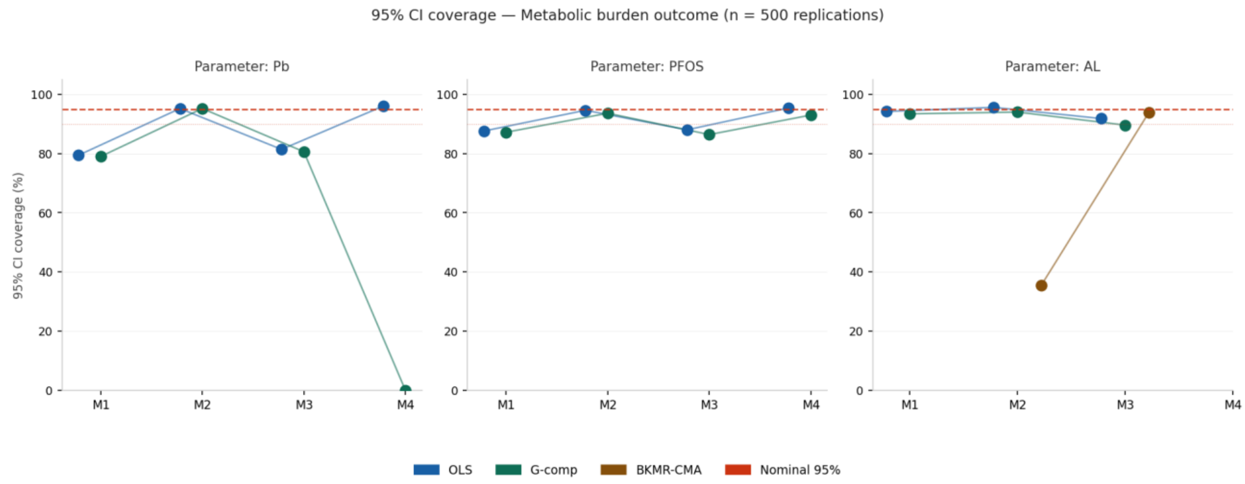


**Figure S3.** 95% CI coverage probability by estimator and mechanism — Metabolic burden outcome, n = 500 replications. Red dashed = nominal 95%; dotted = 90%. BKMR-CMA coverage for AL under M2 is 35.4%, and G-computation Pb coverage under M4 remains 0.0%, consistent with the structural findings in the main CVD analysis.

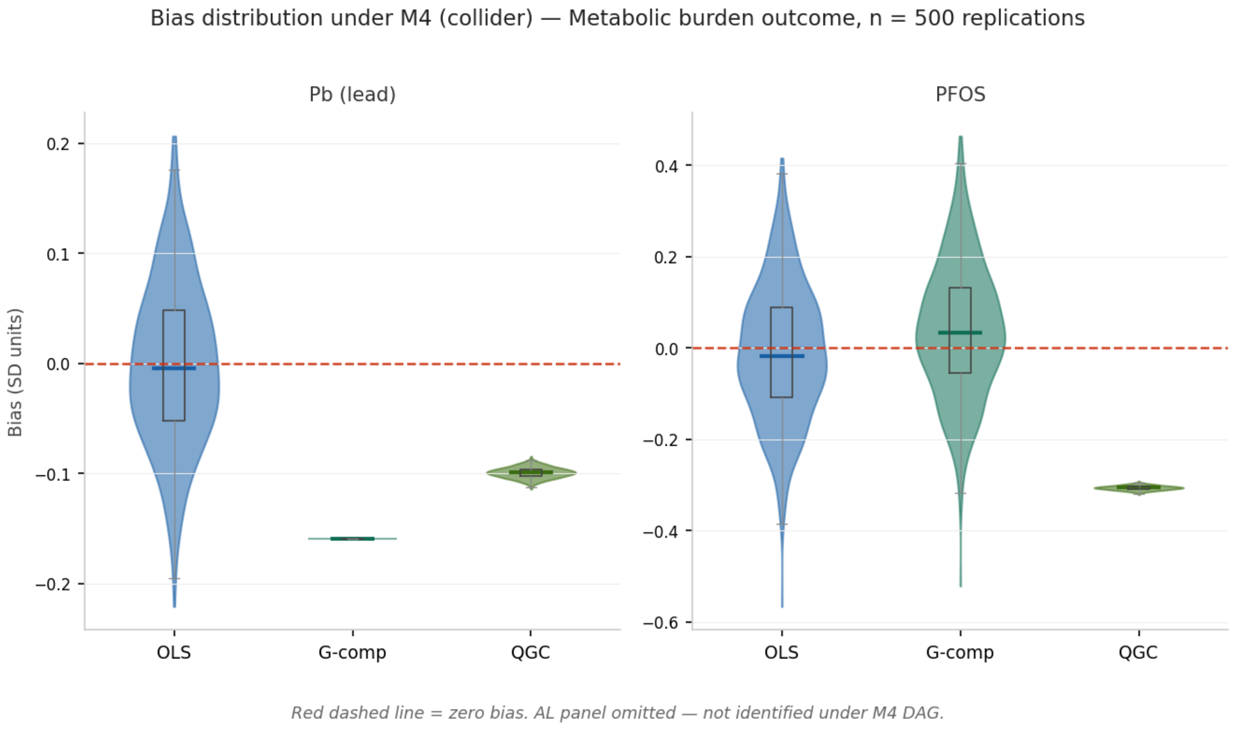


**Figure S4.** Bias distribution under M4 (collider mechanism) — Metabolic burden outcome, n = 500 replications. Violin width = distribution density. G-computation produces a narrow deterministic spike at the non-zero induced bias value; OLS spans a wide distribution centered near zero. AL not shown — not identified under M4.

#### Section B: Supplementary Figures S5–S8: Cancer Latency Outcome (n = 500)

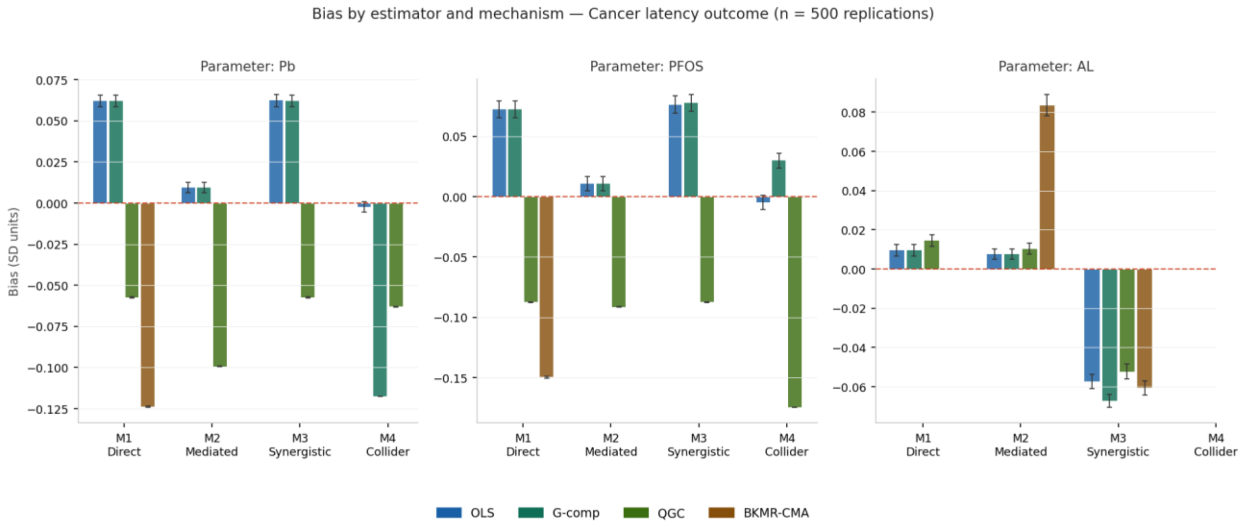


**Figure S5.** Mean bias by estimator and causal mechanism — Cancer latency outcome, n = 500 replications. Effect magnitudes are smaller throughout, reflecting the weaker true effects specified in the cancer DGP (β = 0.05–0.18). The directional pattern of estimator bias is preserved across all mechanisms.

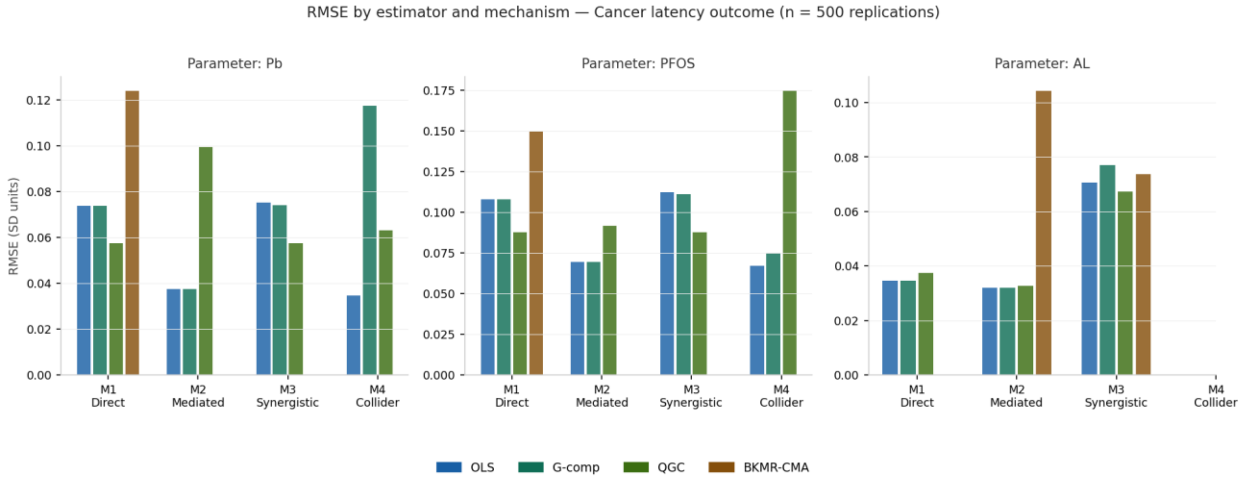


**Figure S6.** RMSE by estimator and mechanism — Cancer latency outcome, n = 500 replications. RMSE values are uniformly smaller than CVD and metabolic outcomes, consistent with smaller true effect sizes. BKMR-CMA RMSE for AL under M2 (+0.104) reflects the same upward bias seen across all three outcomes.

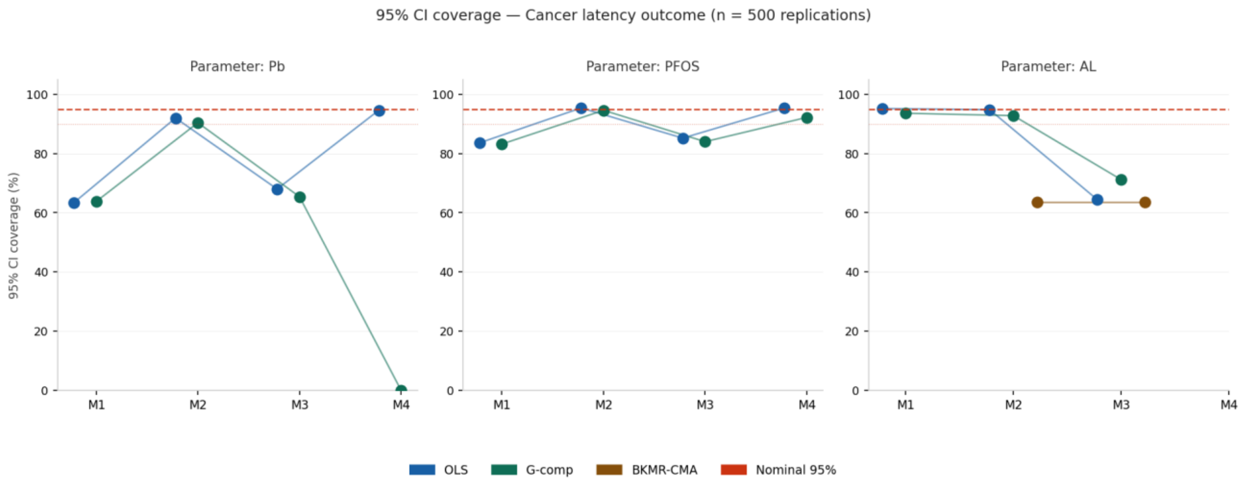


**Figure S7.** 95% CI coverage probability — Cancer latency outcome, n = 500 replications. G-computation Pb coverage under M4 remains 0.0%. BKMR-CMA AL coverage under M2 is 63.6% — higher than CVD (32.8%) due to the weaker exposure-to-AL-to-cancer pathway producing less severe mediation bias. The M3 BKMR-CMA AL coverage advantage (93.6%) is attenuated but persists.

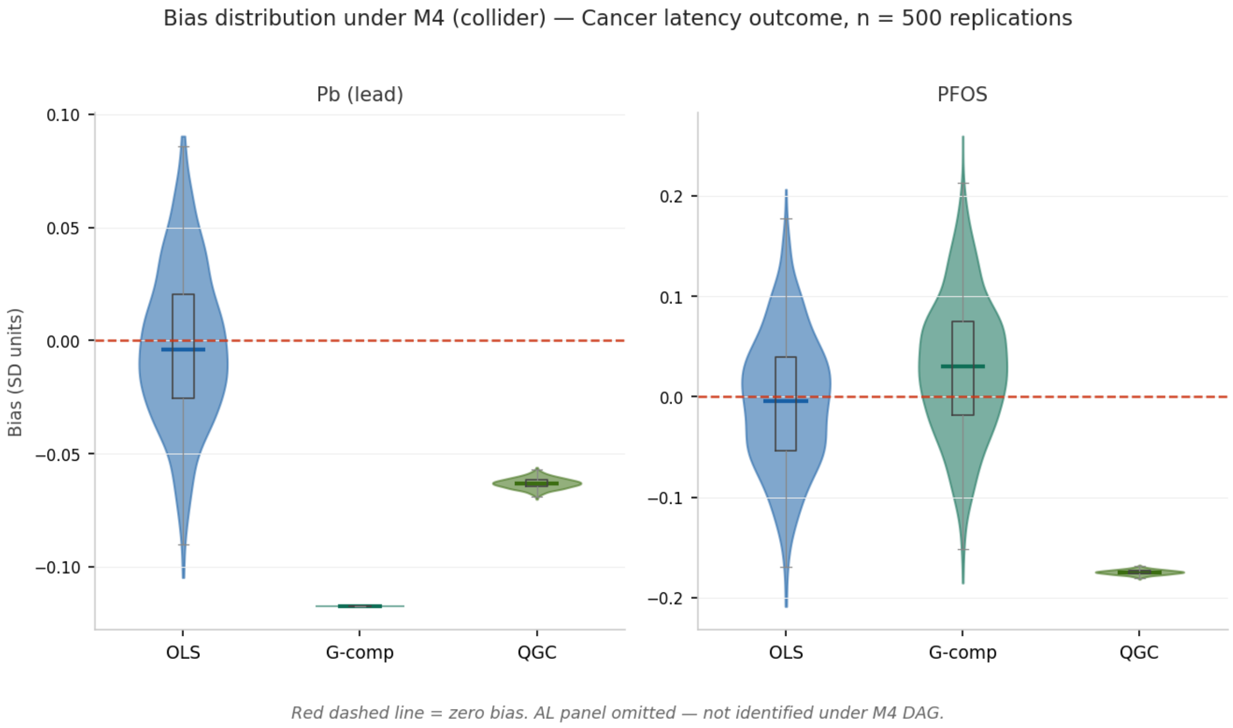


**Figure S8.** Bias distribution under M4 (collider mechanism) — Cancer latency outcome, n = 500 replications. The collider-induced bias distributions are narrower for cancer than for CVD, consistent with smaller path coefficients in the cancer DGP. The structural pattern (narrow G-comp spike, wide OLS distribution) is unchanged.

### Section C: Supplementary Figures S9–S12: CVD Outcome (n = 1,000)

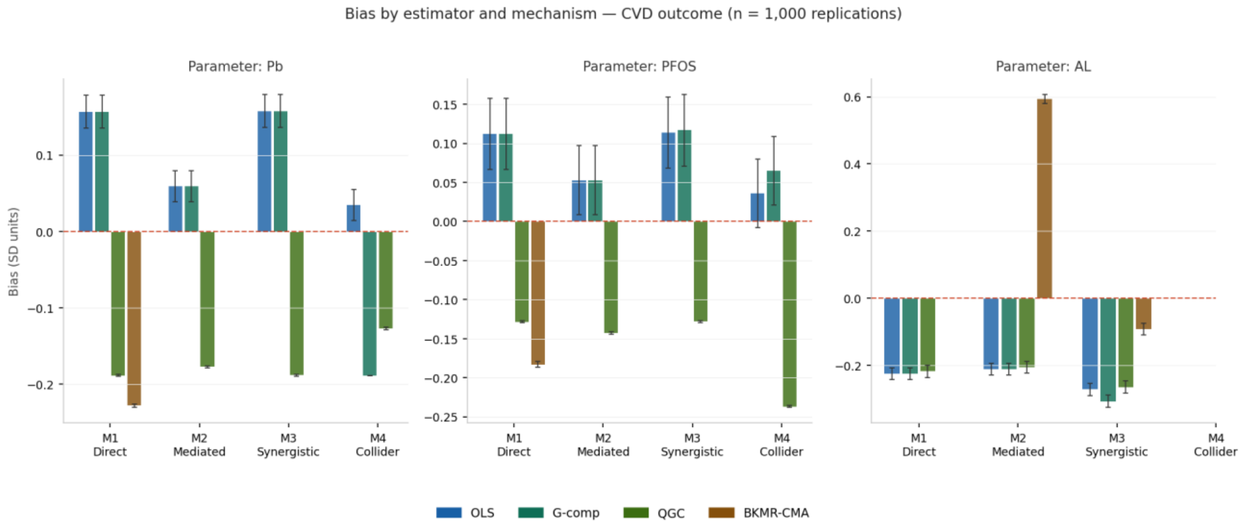


**Figure S9.** Mean bias by estimator and mechanism — CVD outcome, n = 1,000 replications. Bias values are nearly identical to n = 500 results (Figures 2 and S1), confirming that the observed biases are fixed properties of each estimator under each mechanism rather than finite-sample artifacts.

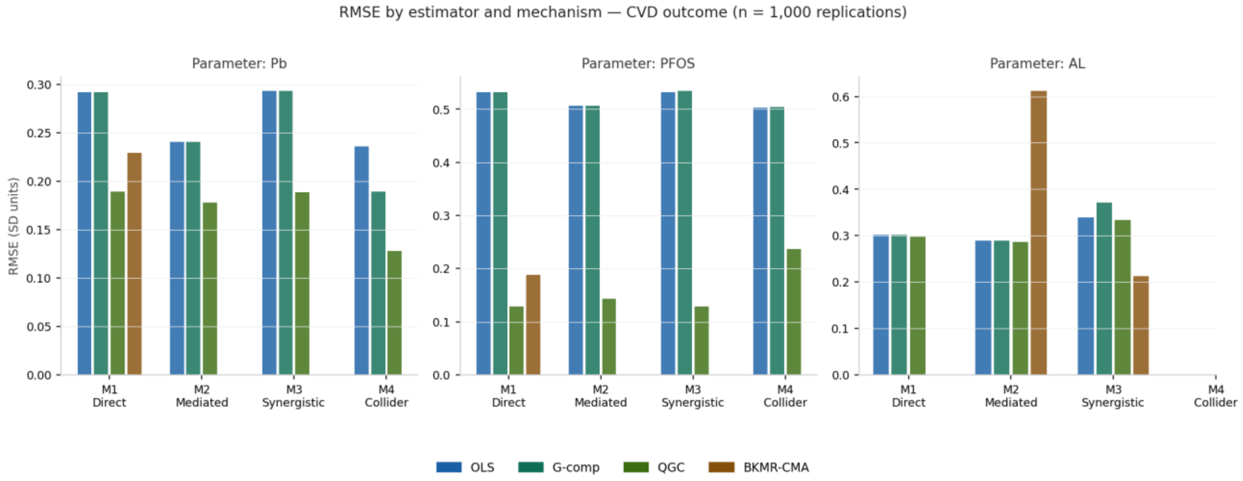


**Figure S10.** RMSE by estimator and mechanism — CVD outcome, n = 1,000 replications. RMSE is visibly reduced relative to n = 500 (Figure 3), particularly for OLS and G-computation on PFOS where variance-driven RMSE decreases with larger sample size. The BKMR-CMA M2 AL spike (RMSE = 0.523 at n = 1,000 vs. 0.611 at n = 500) reflects some improvement but persistent structural bias.

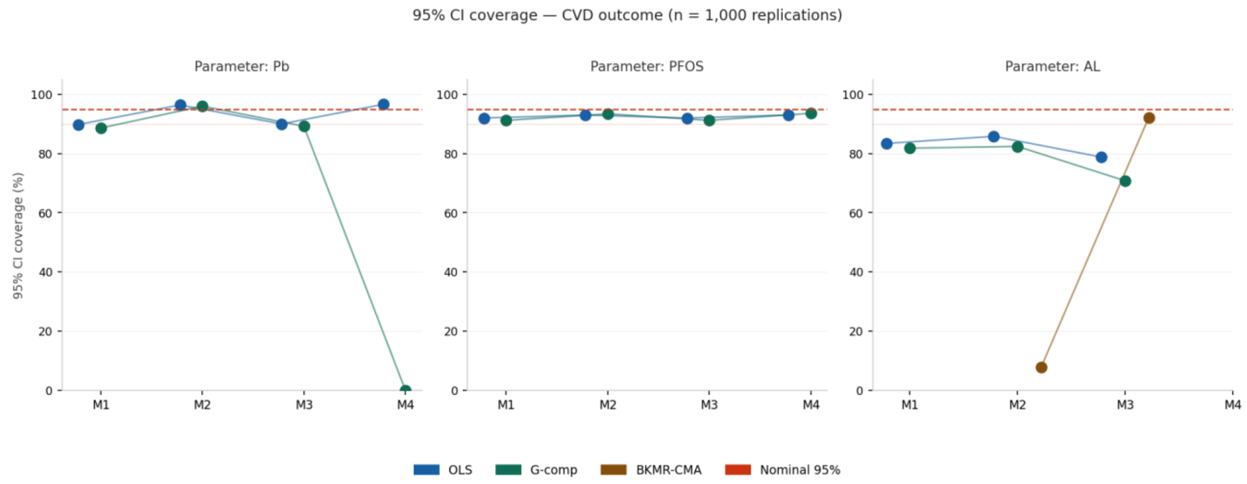


**Figure S11.** 95% CI coverage — CVD outcome, n = 1,000 replications. BKMR-CMA AL coverage under M2 remains severely below nominal (approximately 38%), confirming the coverage failure is structural rather than a small-sample issue. G-computation Pb coverage under M4 remains 0.0%. BKMR-CMA AL coverage under M3 (96.4%) is maintained at nominal levels.

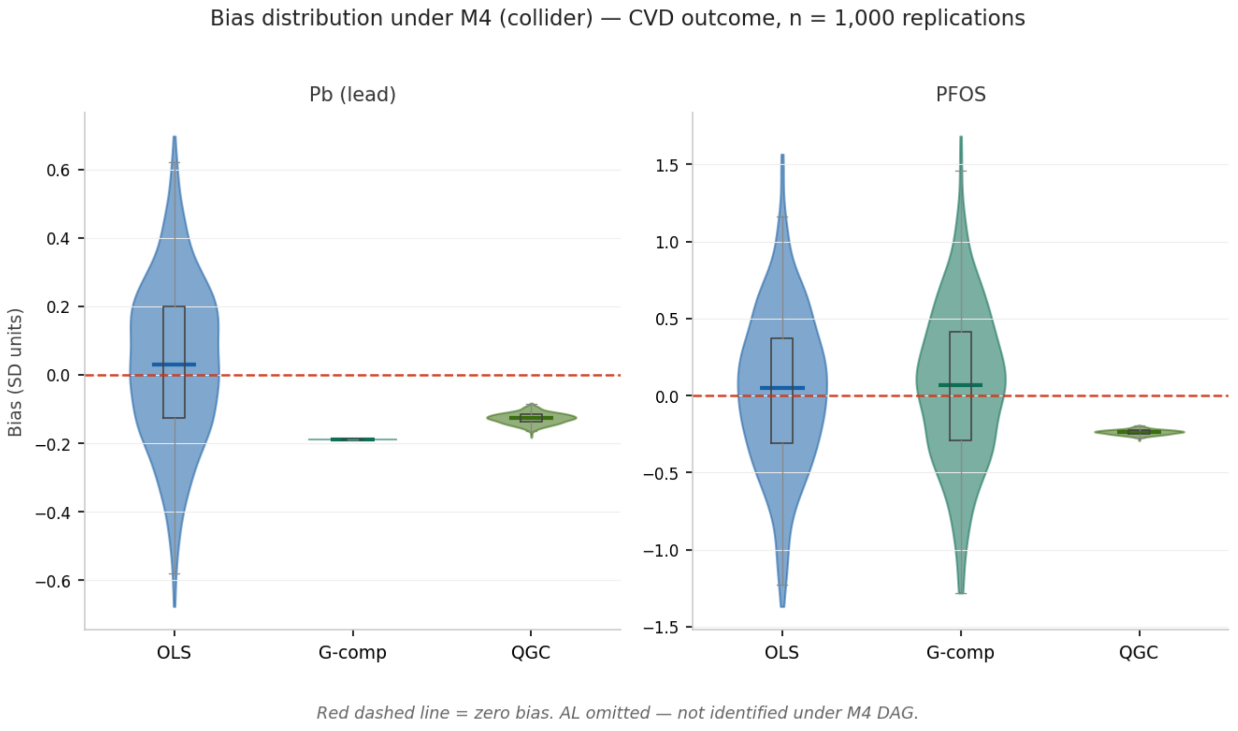


**Figure S12.** Bias distribution under M4 — CVD outcome, n = 1,000 replications. The contrast between the wide OLS violin and the narrow G-computation spike is sharper at n = 1,000 than at n = 500, as increased replication precision tightens the G-computation distribution further around its non-null bias. The structural non-identification interpretation is unchanged.

### Section D: Supplementary Figures S13–S16: Metabolic Burden Outcome (n = 1,000)

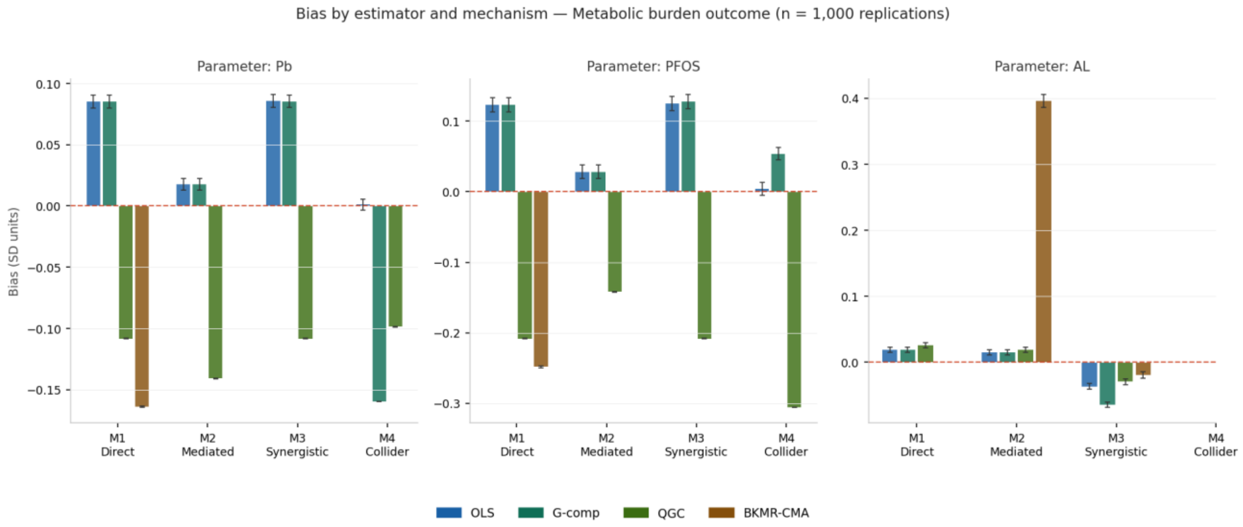


**Figure S13.** Mean bias by estimator and mechanism — Metabolic burden outcome, n = 1,000 replications. Results are consistent with n = 500 (Figure S1); bias direction and relative magnitude are stable across sample sizes, confirming the mechanism-specific bias patterns are not sample-size dependent.

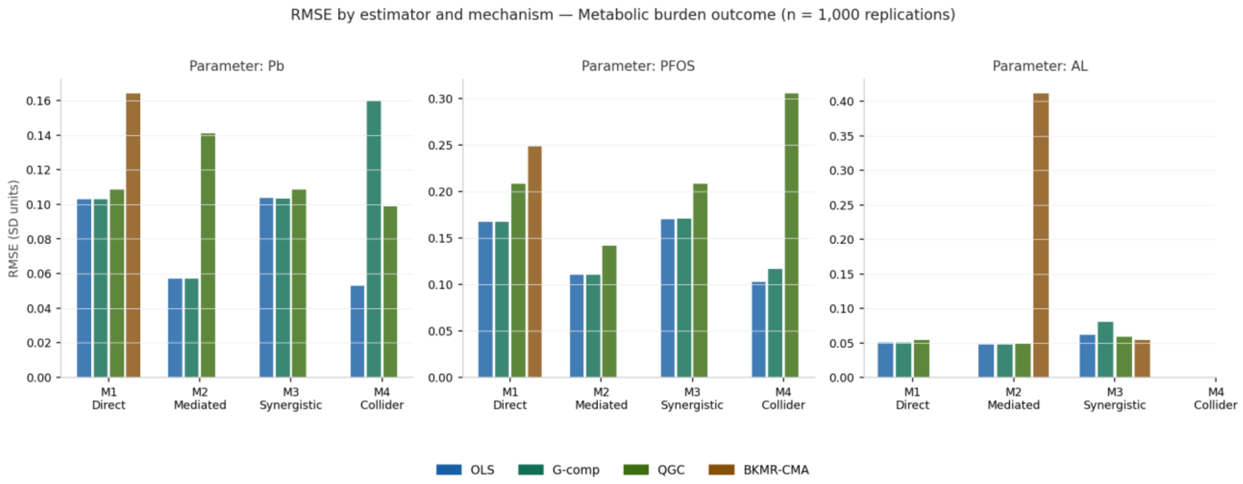


**Figure S14.** RMSE by estimator and mechanism — Metabolic burden outcome, n = 1,000 replications. RMSE reduction from n = 500 to n = 1,000 is most pronounced for OLS and G-computation in the M1 and M2 mechanisms, where variance rather than bias is the dominant contributor to RMSE.

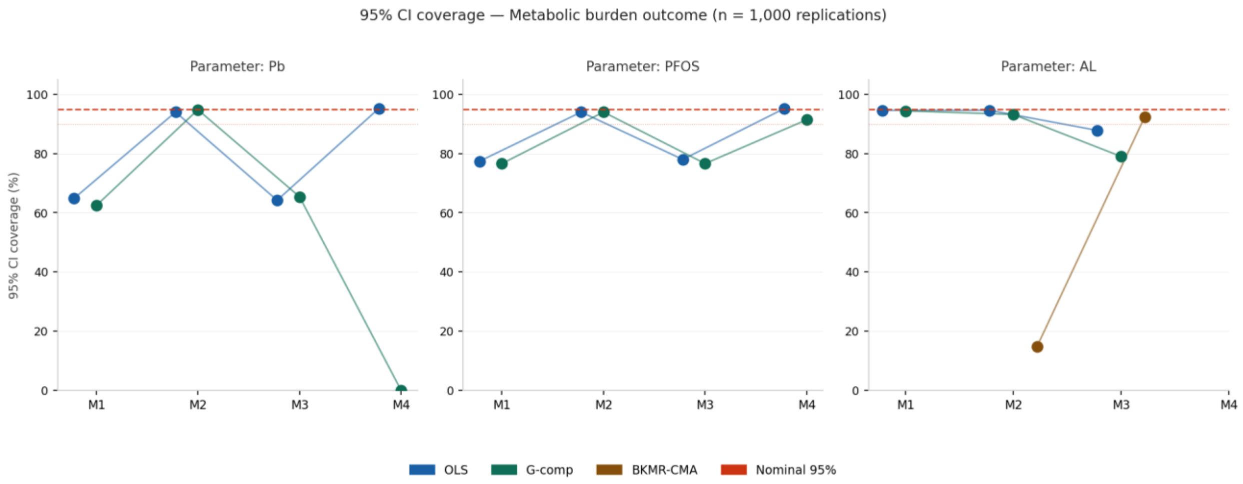


**Figure S15.** 95% CI coverage — Metabolic burden outcome, n = 1,000 replications. The structural coverage failures (G-computation Pb under M4 = 0.0%; BKMR-CMA AL under M2 ≈ 36%) persist at larger sample size, reinforcing that these failures are mechanism-determined rather than variance-driven.

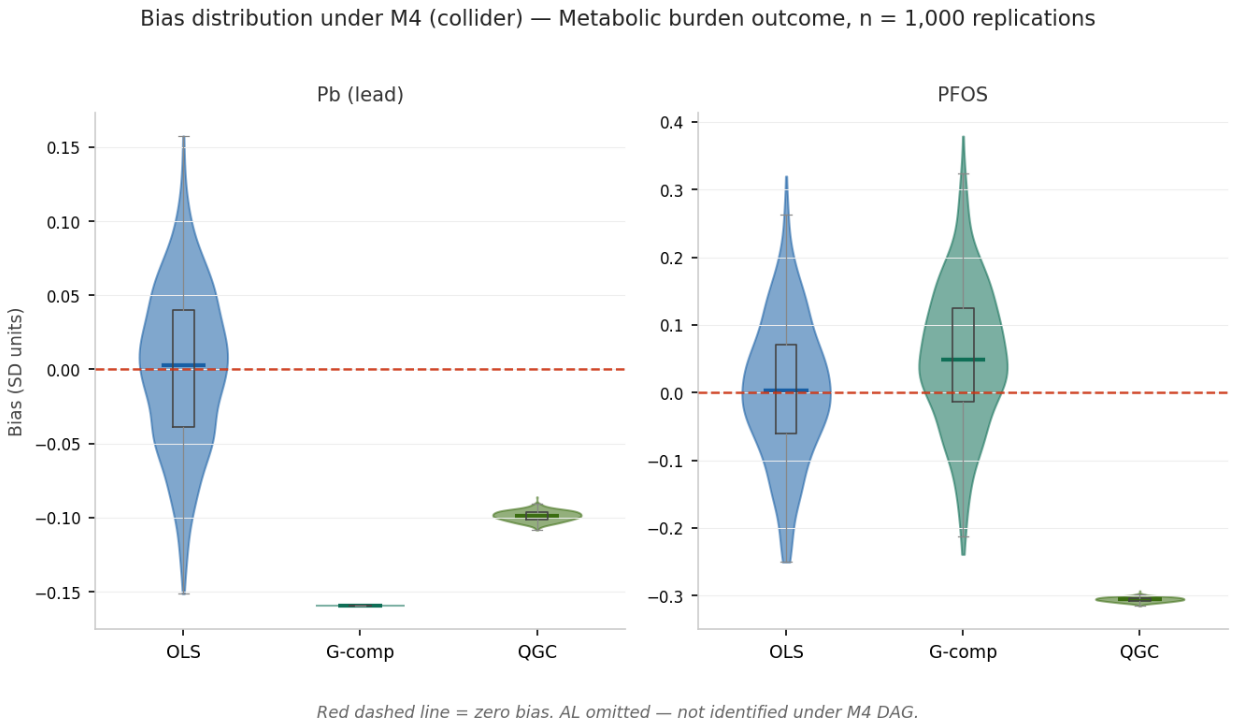


**Figure S16.** Bias distribution under M4 — Metabolic burden outcome, n = 1,000 replications. The collider-induced bias pattern is consistent with n = 500 (Figure S4); the narrowing of all three distributions at larger n is expected and does not affect the causal interpretation of the observed bias patterns.

#### Section E: Supplementary Figures S17–S20: Cancer Latency Outcome (n = 1,000)

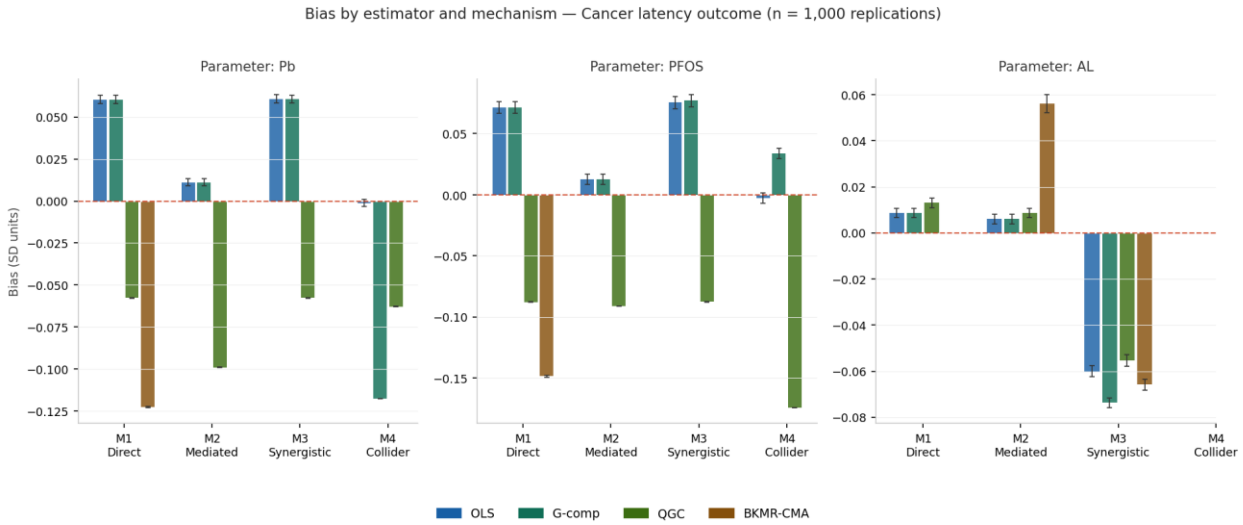


**Figure S17.** Mean bias by estimator and mechanism — Cancer latency outcome, n = 1,000 replications. Effect magnitudes and directions are consistent with n = 500 results (Figure S5). The similarity across sample sizes confirms internal consistency of the simulation.

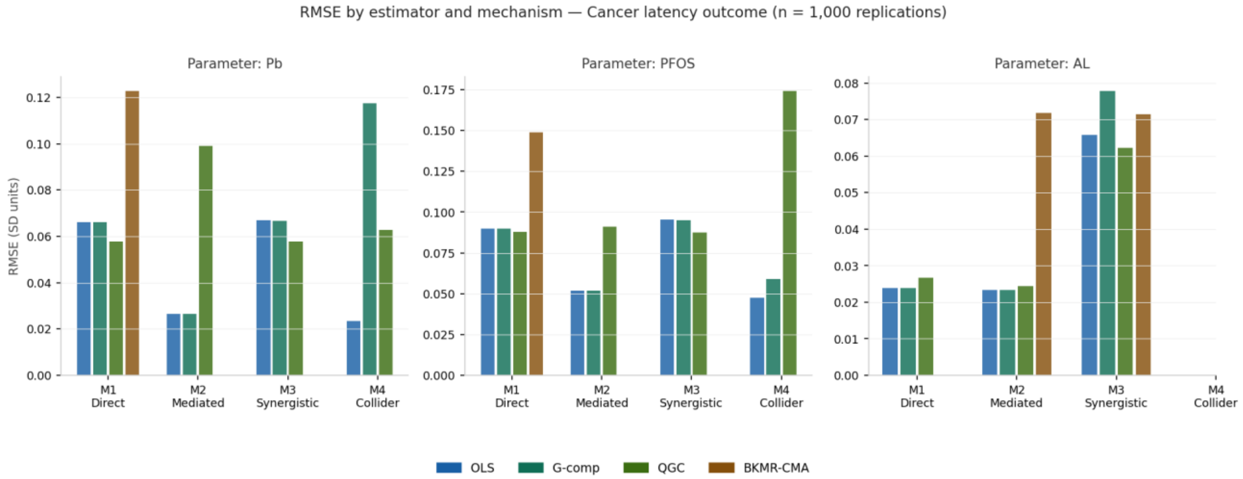


**Figure S18.** RMSE by estimator and mechanism — Cancer latency outcome, n = 1,000 replications. RMSE values decrease uniformly from n = 500 to n = 1,000 for all estimators and mechanisms. The rank ordering of estimators within each mechanism-parameter cell is preserved across sample sizes.

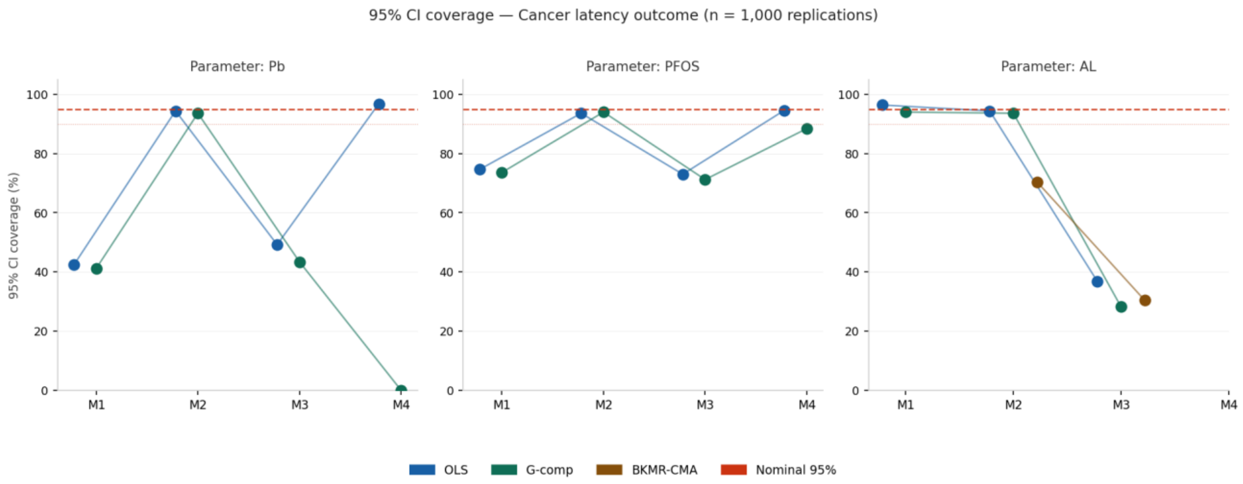


**Figure S19.** 95% CI coverage — Cancer latency outcome, n = 1,000 replications. G-computation Pb coverage under M4 remains 0.0%. BKMR-CMA AL coverage under M2 improves modestly to approximately 68% at n = 1,000 (from 63.6% at n = 500), consistent with small variance-driven recovery as MCMC chains stabilize. The coverage remains far below nominal.

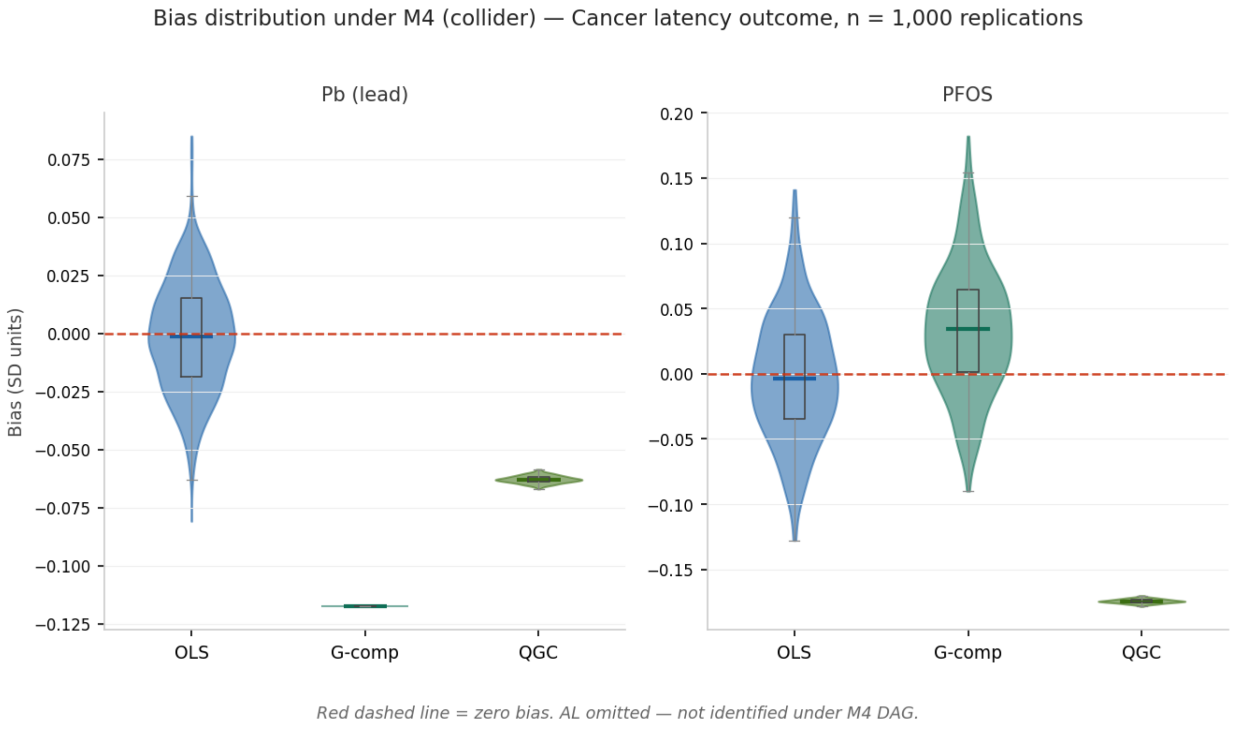


**Figure S20.** Bias distribution under M4 — Cancer latency outcome, n = 1,000 replications. Distributions are tighter at larger n; the structural collider-bias interpretation is unchanged. This figure, together with Figures S4, S8, S12, and S16, demonstrates that the collider bias findings are consistent across all three outcomes and both sample sizes evaluated.

#### References

1. Centers for Disease Control and Prevention, National Center for Health Statistics. National Health and Nutrition Examination Survey Data 2015–2018. Hyattsville, MD: U.S. Department of Health and Human Services; 2018. Available at: https://www.cdc.gov/nchs/nhanes/. Accessed May 21, 2026.

2. Agency for Toxic Substances and Disease Registry. Toxicological Profile for Perfluoroalkyls. Atlanta, GA: U.S. Department of Health and Human Services; 2021. Available at: https://www.atsdr.cdc.gov/toxprofiles/tp200.pdf. Accessed May 21, 2026.

3. Ruckart PZ, Bove FJ, Maslia M. Evaluation of exposure to contaminated drinking water and specific birth defects and childhood cancers at Marine Corps Base Camp Lejeune, North Carolina: a case-control study. Environ Health. 2013;12:104.

4. Institute of Medicine. Gulf War and Health, Volume 9: Long-Term Effects of Blast Exposures. Washington, DC: The National Academies Press; 2014.

5. Kok BC, Herrell RK, Thomas JL, Hoge CW. Posttraumatic stress disorder associated with combat service in Iraq or Afghanistan: reconciling prevalence differences between studies. J Nerv Ment Dis. 2012;200:444–450.

6. Stevelink SAM, Jones M, Hull L, et al. Post-traumatic stress disorder, depression and anxiety in UK armed forces personnel: prevalence rates and relating to mental health care. BMJ. 2018;362:k2948.

7. Brailey K, Vasterling JJ, Proctor SP, Constans JI, Friedman MJ. PTSD symptoms, life events, and unit cohesion in U.S. soldiers: baseline findings from the neurocognition deployment health study. J Trauma Stress. 2007;20:495–503.

8. Morgan CA 3rd, Wang S, Mason J, et al. Hormone profiles in humans experiencing military survival training. Biol Psychiatry. 2000;47:891–901.

9. D'Agostino RB Sr, Vasan RS, Pencina MJ, et al. General cardiovascular risk profile for use in primary care: the Framingham Heart Study. Circulation. 2008;117:743–753.

10. Juster RP, McEwen BS, Lupien SJ. Allostatic load biomarkers of chronic stress and impact on health and cognition. Neurosci Biobehav Rev. 2010;35:2–16.

11. Seeman TE, Epel E, Gruenewald T, Karlamangla A, McEwen BS. Socio-economic differentials in peripheral biology: cumulative allostatic load. Ann N Y Acad Sci. 2010;1186:223–239.

12. Weisskopf MG, Jain N, Nie H, et al. A prospective study of bone lead concentration and death from all causes, cardiovascular diseases, and cancer in the normative aging study. Circulation. 2009;120:1056–1064.

13. Tellez-Plaza M, Guallar E, Howard BV, et al. Cadmium exposure and incident cardiovascular disease. Epidemiology. 2013;24:421–429.

14. Virtanen JK, Rissanen TH, Voutilainen S, Tuomainen TP. Mercury as a risk factor for cardiovascular diseases. J Nutr Biochem. 2007;18:75–85.

15. Fitz-Simon N, Fletcher T, Luster MI, et al. Reductions in serum lipids with a 4-year decline in serum perfluorooctanoic acid and perfluorooctanesulfonic acid concentrations in the Arnsberg cohort. Am J Epidemiol. 2013;178:1443–1454.

16. Lind PM, van Bavel B, Salihovic S, Lind L. Circulating levels of persistent organic pollutants (POPs) and carotid atherosclerosis in the elderly. Environ Health Perspect. 2012;120:38–43.

17. Gallo V, Leonardi G, Genser B, et al. Serum perfluorooctanoate (PFOA) and perfluorooctane sulfonate (PFOS) concentrations and liver function biomarkers in a population with elevated PFOA exposure. Environ Health Perspect. 2012;120:655–660.

18. Satarug S, Moore MR. Adverse health effects of chronic exposure to low-level cadmium in foodstuffs and cigarette smoke. Environ Health Perspect. 2004;112:1099–1103.

19. International Agency for Research on Cancer. IARC Monographs on the Evaluation of Carcinogenic Risks to Humans, Volume 100C: Arsenic, Metals, Fibres and Dusts. Lyon, France: IARC; 2012.

20. International Agency for Research on Cancer. IARC Monographs on the Evaluation of Carcinogenic Risks to Humans, Volume 107: Polychlorinated Biphenyls and Polybrominated Biphenyls. Lyon, France: IARC; 2016.

21. Barry V, Winquist A, Steenland K. Perfluorooctanoic acid (PFOA) exposures and incident cancers among adults living near a chemical plant. Environ Health Perspect. 2013;121:1313–1318.

22. Grandjean P, Andersen EW, Budtz-Jørgensen E, et al. Serum vaccine antibody concentrations in children exposed to perfluorinated compounds. JAMA. 2012;307:391–397.

23. Navas-Acien A, Guallar E, Silbergeld EK, Rothenberg SJ. Lead exposure and cardiovascular disease: a systematic review. Environ Health Perspect. 2007;115:472–482.

24. Hamra GB, Guha N, Cohen A, et al. Outdoor particulate matter exposure and lung cancer: a systematic review and meta-analysis. Environ Health Perspect. 2014;122:906–911.

25. Bobb JF, Valeri L, Claus Henn B, et al. Bayesian kernel machine regression for estimating the health effects of multi-pollutant mixtures. Biostatistics. 2015;16:493–508.

26. Keil AP, Buckley JP, O'Brien KM, Ferguson KK, Zhao S, White AJ. A quantile-based g-computation approach to addressing the effects of exposure mixtures. Environ Health Perspect. 2020;128:047004.

27. Morris TP, White IR, Crowther MJ. Using simulation studies to evaluate statistical methods. Stat Med. 2019;38:2074–2102.
